# Biallelic variants in *POPDC2* cause a novel autosomal recessive syndrome presenting with cardiac conduction defects and variable hypertrophic cardiomyopathy

**DOI:** 10.1101/2024.07.04.24309755

**Authors:** Michele Nicastro, Alexa M.C. Vermeer, Pieter G. Postema, Rafik Tadros, Forrest Z. Bowling, Hildur M. Aegisdottir, Vinicius Tragante, Lukas Mach, Alex V. Postma, Elisabeth M. Lodder, Karel van Duijvenboden, Rob Zwart, Leander Beekman, Lingshuang Wu, Paul A. van der Zwaag, Mariëlle Alders, Mona Allouba, Yasmine Aguib, J. Luis Santomel, David de Una, Lorenzo Monserrat, Antonio M. A. Miranda, Kazumasa Kanemaru, James Cranley, Ingeborg E. van Zeggeren, Eleonora M.A. Aronica, Michela Ripolone, Simona Zanotti, Gardar Sveinbjornsson, Erna V. Ivarsdottir, Hilma Hólm, Daníel F. Guðbjartsson, Ástrós Th. Skúladóttir, Kári Stefánsson, Lincoln Nadauld, Kirk U. Knowlton, Sisse Rye Ostrowski, Erik Sørensen, Ole Birger Vesterager Pedersen, Jonas Ghouse, Søren Rand, Henning Bundgaard, Henrik Ullum, Christian Erikstrup, Bitten Aagaard, Mie Topholm Bruun, Mette Christiansen, Henrik K. Jensen, Deanna Alexis Carere, Christopher T Cummings, Kristen Fishler, Pernille Mathiesen Tøring, Klaus Brusgaard, Trine Maxel Juul, Lotte Saaby, Bo Gregers Winkel, Jens Mogensen, Francesco Fortunato, Giacomo Pietro Comi, Dario Ronchi, J. Peter van Tintelen, Michela Noseda, Michael V. Airola, Imke Christiaans, Arthur A.M. Wilde, Ronald Wilders, Sally-Ann Clur, Arie O. Verkerk, Connie R. Bezzina, Najim Lahrouchi

**Affiliations:** Amsterdam UMC, University of Amsterdam, Heart Center, Department of Clinical and Experimental Cardiology, Amsterdam Cardiovascular Sciences, Amsterdam, The Netherlands; Member of the European Reference Network for rare, low prevalence and complex diseases of the heart: ERN GUARD-Heart; Department of Human Genetics, Amsterdam UMC, University of Amsterdam, Amsterdam, The Netherlands; Cardiovascular Genetics Center, Montreal Heart Institute and Faculty of Medicine, Université de Montréal, Montreal, Canada; Department of Biochemistry and Cell Biology, Stony Brook University, Stony Brook, NY, USA; deCODE genetics/Amgen, Inc., Reykjavik, Iceland; Faculty of Medicine, University of Iceland, Reykjavik, Iceland; National Heart and Lung Institute, Imperial College London, UK; Royal Brompton Hospital, London, UK; Department of Medical Biology, Amsterdam UMC, University of Amsterdam, Amsterdam, The Netherlands; Department of Genetics, University of Groningen, University Medical Center Groningen, Groningen, The Netherlands; Aswan Heart Centre, Magdi Yacoub Foundation, Egypt; NHLI, Imperial College, London, UK; Health in Code, A Coruña, Spain; Medical Department, Dilemma Solutions SL. Cardiovascular Research Group A Coruña University; Wellcome Sanger Institute, Wellcome Genome Campus, Hinxton, Cambridge, UK; Department of Neurology, Amsterdam UMC, Amsterdam Neuroscience, University of Amsterdam, Amsterdam, The Netherlands; Department of Neuropathology, Amsterdam UMC location University of Amsterdam, Amsterdam Neuroscience, Amsterdam, The Netherlands; Fondazione IRCCS Ca’ Granda Ospedale Maggiore Policlinico, Neuromuscular and Rare Disease Unit, Milan, Italy; Intermountain Healthcare, St. George, UT, USA; Intermountain Healthcare, Salt Lake City, UT, USA; Department of Clinical Immunology, Copenhagen University Hospital, Rigshospitalet, Copenhagen, Denmark; Department of Clinical Medicine, Faculty of Health and Medical Sciences, University of Copenhagen, Copenhagen, Denmark; Department of Clinical Immunology, Zealand University Hospital, Køge, Denmark; Department of Cardiology, Copenhagen University Hospital, Rigshospitalet, Copenhagen, Denmark; Statens Serum Institut, Copenhagen, Denmark; Department of Clinical Immunology, Aarhus University Hospital, Aarhus, Denmark; Department of Clinical Medicine, Health, Aarhus University, Denmark; Department of Clinical Immunology, Aalborg University Hospital, Aalborg, Denmark; Clinical Immunology Research Unit, Department of Clinical Immunology, Odense University Hospital, Odense, Denmark; Department of Molecular Medicine, Aarhus University Hospital, Denmark; Department of Cardiology, Aarhus University Hospital, Denmark; GeneDx, Gaithersburg, Maryland, USA; Department of Pediatrics, Division of Genetics, University of Nebraska Medical Center, Omaha, NE, USA; Children’s Nebraska, Department of Genetic Medicine, Omaha, NE, USA; Department of Clinical Genetics, Odense University Hospital, Odense, Denmark; Department of Clinical Genetics, Lillebaelt Hospital, Vejle, Denmark; Institute for Regional Health Services, University of Southern Denmark, Campusvej 55, 5230 Odense, Denmark; Department of Cardiology, Odense University Hospital, Odense, Denmark; Department of Cardiology, Aalborg University Hospital, Denmark; Dino Ferrari Center, Department of Pathophysiology and Transplantation, University of Milan, Milan, Italy; IRCCS Fondazione Cà Granda Ospedale Maggiore Policlinico, Neurology Unit, Milan, Italy; Department of Genetics, University Medical Center Utrecht, Utrecht University, Utrecht, The Netherlands; Imperial British Heart Foundation Center for Research Excellence; Department of Pediatric Cardiology, Emma Children’s Hospital, Amsterdam UMC, University of Amsterdam, Amsterdam, The Netherlands

## Abstract

*POPDC2* encodes for the Popeye domain-containing protein 2 which has an important role in cardiac pacemaking and conduction, due in part to its cAMP-dependent binding and regulation of TREK-1 potassium channels. Loss of *Popdc2* in mice results in sinus pauses and bradycardia and morpholino knockdown of *popdc2 in* zebrafish results in atrioventricular (AV) block. We identified bi-allelic variants in *POPDC2* in 4 families that presented with a phenotypic spectrum consisting of sinus node dysfunction, AV conduction defects and hypertrophic cardiomyopathy. Using homology modelling we show that the identified *POPDC2* variants are predicted to diminish the ability of POPDC2 to bind cAMP. In *in vitro* electrophysiological studies we demonstrated that, while co-expression of wild-type POPDC2 with TREK-1 increased TREK-1 current density, POPDC2 variants found in the patients failed to increase TREK-1 current density. While patient muscle biopsy did not show clear myopathic disease, it showed significant reduction of the expression of both POPDC1 and POPDC2, suggesting that stability and/or membrane trafficking of the POPDC1–POPDC2 complex is impaired by pathogenic variants in any of the two proteins. Single-cell RNA sequencing from human hearts demonstrated that co-expression of POPDC1 and 2 was most prevalent in AV node, AV node pacemaker and AV bundle cells. Sinoatrial node cells expressed POPDC2 abundantly, but expression of POPDC1 was sparse. Together, these results concur with predisposition to AV node disease in humans with loss-of-function variants in *POPDC1* and POPDC2 and presence of sinus node disease in *POPDC2,* but not in *POPDC1* related disease in human. Using population-level genetic data of more than 1 million individuals we showed that none of the familial variants were associated with clinical outcomes in heterozygous state, suggesting that heterozygous family members are unlikely to develop clinical manifestations and therefore might not necessitate clinical follow-up. Our findings provide evidence for *POPDC2* as the cause of a novel Mendelian autosomal recessive cardiac syndrome, consistent with previous work showing that mice and zebrafish deficient in functional *POPDC2* display sinus and AV node dysfunction.

**GRAPHICAL ABSTRACT:** 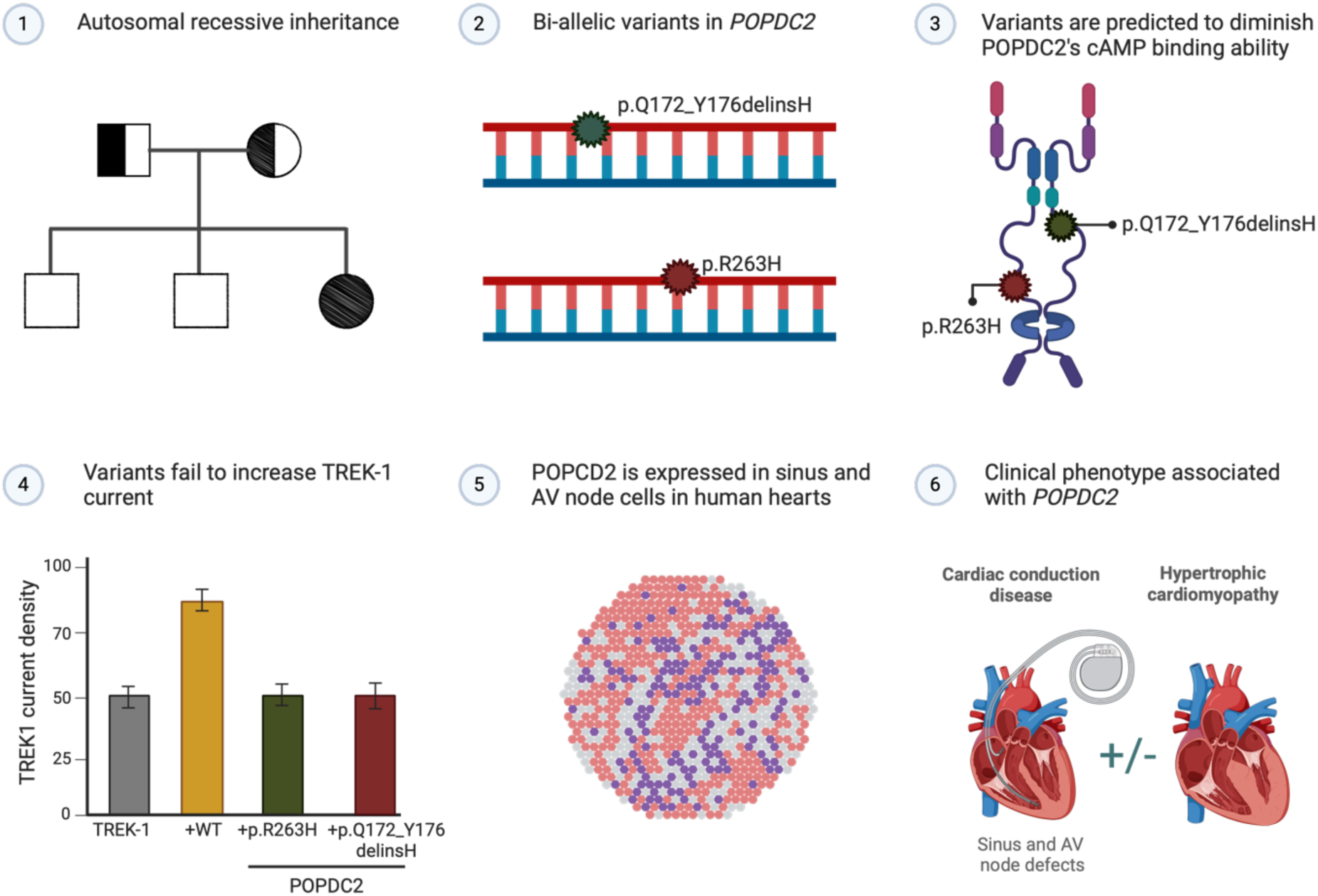

## INTRODUCTION

The rhythmic contraction of the heart is orchestrated by the cardiac pacemaker and conduction system.^1^ Electrical activity in the heart arises in the sinus node, located in the right atrium near the entrance of the superior vena cava. The electrical impulse then spreads through the atria to the atrioventricular (AV) node and is subsequently propagated through the bundle of His and bundle branches, to the Purkinje fibers from where it spreads throughout the ventricles.

Cardiac conduction defects (CCDs) are primarily the consequence of age-related degeneration, structural heart disease or post-operative complications.^2^ Presentation of CCDs in the young should raise suspicion of a genetic disorder. Rare variants in genes encoding cardiac ion channels (e.g. *SCN5A*, *TRPM4* and *HCN4*), transcription factors (e.g. *TBX5*, *NKX2-5*), constituents of the inner nuclear membrane (e.g. *LMNA*, *EMD*), gap junction proteins (e.g. *GJC1*) and others (e.g. *GNB5*, *TNNI3K*, *PRKAG2*) have been implicated in inherited CCD presenting in isolation or in presence of other cardiac or extracardiac features.^2^ Yet, many patients with early onset CCD remain genetically unexplained.

Recessive variants in *POPDC1* (also known as *BVES*), encoding the Popeye domain-containing protein 1, were associated with muscular dystrophy and AV block.^3,4^ In mice, knockout of *Popdc1* or *Popdc2* resulted in stress-induced sinus pauses and sinus bradycardia.^5^ In zebrafish, morpholino knockdown of *popdc1* or *popdc2,* resulted in second-degree AV block and bradycardia.^3,6^ The TWIK-related potassium channel 1 (TREK-1, encoded by *KCNK2*) is an established interacting protein of POPDC2^5^ and co-expression of POPDC2 and TREK-1 has been shown to result in a 2-fold higher TREK-1 current in comparison to expression of TREK-1 alone.^5^ Here, we provide evidence for homozygous loss-of-function variants in *POPDC2* as the cause of a novel autosomal recessive syndrome in four families, consisting of a phenotypic spectrum including sinus node disease and AV conduction defects with variable hypertrophic cardiomyopathy (HCM).

## MATERIAL AND METHODS

### Case recruitment and DNA sequencing

Family A was referred to the Department of Human Genetics for genetic testing and counselling for CCD and HCM. To follow up on the findings from exome sequencing in this family, we studied 78 patients that were diagnosed with a similar clinical presentation as Family A (i.e., CCDs and HCM, cohorts 1-3). In addition, we studied 96 HCM patients without CCDs (**cohort 4**). In all 174 patients, genetic testing had ruled out causative variants in established arrhythmia and/or cardiomyopathy genes. Families C and D were identified via a genetic and phenotypic match through DECIPHER^7^ and GeneMatcher^8^, respectively. The study protocol was approved by the local Institutional Review Board and signed informed consent was obtained from the patients or their parents. Details on case recruitment and DNA sequencing methods for each family can be found in the **Supplemental Methods and Supplemental Table 1**. To ensure the privacy of the patients and their families; (1) ages are presented as non-overlapping age ranges (i.e., 0-5, 6-10, 11-15 years), (2) pedigrees were modified, (3) information related to ancestry/country or origin/nationality are not reported and (4) clinical descriptions were minimized. The corresponding author should be contacted to request access to the full data.

### Homology modeling of *POPDC2*

Two homology models were generated using either SWISS-MODEL^9^ or AlphaFold2 Multimer.^10^ The SWISS-MODEL web server used a cAMP-Regulatory Protein from *Yersinia pestis* (6DT4) as a template to generate the homology model for the Popeye domain of POPDC2. A dimer of full-length POPDC2 was generated using AlphaFold Multimer^10^ and the intrinsically disordered C-terminal residues 275-364 were deleted to simplify figure presentation. A final homology model of POPDC2 was created by replacing residues 128-213 of the AlphaFold Multimer model with residues 128-213 of the SWISS-cAMP model, after superimposing the individual Popeye domains.

AlphaMissense^11^ was used to generate the predicted pathogenicity of single amino acid substitutions and deleted regions. AlphaMissense uses language modeling to understand amino acid distributions based on sequence context, then incorporates structural information using an AlphaFold derived system to consider a protein’s three-dimensional form when assessing a variants’ impact. It also utilizes weak labels from population frequency data to refine predictions without human biases. Using these models we evaluated the consequence of POPDC2 variants found in Families A-D and five variants that occurred homozygously in individuals from the Genome Aggregation Database (gnomAD)^12^, which are not expected to cause disease (**Supplemental Table 2**)

### Evaluation of muscle biopsy

#### Histological analysis

Tissue specimens (m. vastus lateralis) were frozen in isopentane-cooled liquid nitrogen and processed according to standard techniques.^13^ For histological analysis, 8 µm-thick cryosections were picked and processed for routine staining with Haematoxylin and Eosin (H&E) and Modified Gomori Trichrome (MGT).

#### Muscle section immunofluorescence staining

Immunofluorescence staining was performed on 8 μm-thick cryostat sections using the following antibodies: Anti-POPDC1 (BVES) (1:100, rabbit polyclonal; Sigma Life Science), Anti-POPDC2 (1:200, rabbit polyclonal; Sigma Life Science) and Anti-Caveolin-3 (1:1000, mouse monoclonal; BD Transduction, East Rutherford, NJ, USA). Slides were then incubated with the appropriate secondary antibody conjugated to Alexa 488 or Alexa 568 (1:2000, Invitrogen Life Technologies). Sections were mounted in Vectashield anti-fade mounting medium containing DAPI (Vector Laboratorie, Burlingame, CA, USA). Image fields were acquired using optical microscope Leica DM4000B equipped with camera (DFC420C).

#### Electron microscopy

For ultrastructural examination, a small part of muscle sample was fixed in 2.5% glutaraldehyde (pH 7.4), post fixed in 2% osmium tetroxide and then, after dehydration in a graded series of ethanols, embedded in Epon’s resin. Finally, ultrathin sections were stained with lead citrate and uranyl acetate replacement stain and examined with Zeiss EM109 transmission electron microscope.

#### Protein studies in muscle specimens

Protein samples were extracted in urea-SDS and protein content evaluated by Lowry method. Thirty micrograms of protein treated with 4% β-mercaptoethanol were run on a 10% polyacrylamide gel and then transferred on a nitrocellulose membrane, saturated with Intercept Blocking Buffer PBS (Licor) for one hour at room temperature. The membrane was incubated at 4°C overnight in a blocking solution containing primary antibodies: anti-POPDC2 (Sigma HPA024255, 1:300), anti-POPDC1 (Sigma HPA014788, 1:300), anti-actinin (Sigma A7811; 1:5500). The next day, the membrane was washed five times with PBS-T (Tween 20, 0.08%) and incubated at room temperature for one hour with the appropriate secondary antibodies in blocking solution (IRDye Licor). The bands corresponding to the proteins of interest were visualized on a Licor Odissey XF imaging system.

### POPDC gene expression profiles in single-nucleus and spatial transcriptomics of human hearts

Single nuclei RNA sequencing data and Visium spatial gene expression data were obtained from a previously published study.^14^ Annotated, log-normalised count matrices for both modalities were downloaded and specifically analyzed for expression of POPDC1, 2 and 3 using Scanpy package for Python run in Jupyter Notebook. Original histological annotation of tissue sections was used. The cell state annotation was adapted from the original study.^14^ All atrial cardiomyocytes were pooled in 1 category, all ventricular cardiomyocytes were pooled together. From the conduction system cells, sinoatrial node pacemaker cells (SAN P cell) and Purkinje cells are shown separately; due to low cell numbers, atrioventricular node pacemaker and bundle cells (AVN P and bundle cells) are shown together.

### POPDC gene expression profiles in single cell RNA sequencing data in mice

Sinus node and AV node/His single cell RNA-sequencing data from mice were obtained from a previously published study.^15^ Using these data, t-distributed stochastic neighbor embedding (t-SNE) maps with a perplexity of 50 were generated on the R2 environment Genomics Analysis and Visualization Platform (http://r2.amc.nl).^16^ The cells were subsequently clustered into different populations using the t-SNE DBSCAN tool. Sentinel gene expression was used to characterize the different clusters (e.g. sinus node cells, expressing higher levels of Tbx3, Isl1 and Hcn4). Thereafter, POPDC protein gene expression intensities were plotted on the t-SNE maps to identify their expression profiles across the present tissue clusters.

### cDNA constructs and mutagenesis

hTREK-1a cloned in pIRES2-EGFP was obtained from Drs. Delphine Bichet and Florian Lesage (Université Nice Sophia Antipolis, France). Full-length *POPDC2* cDNA sequences (NM_001308333-hg19; wild-type, c.516_528C:p.Q172_Y176delinsH and c.G788A:p.R263H) were synthesized, cloned into pBluescript IISK+ (GeneCust, Boynes, France) and subsequently subcloned into pIRES-GFP (pCGI).

### Cell preparation and expression

The effect of mutant POPDC2 on the TREK-1 current was studied in HEK-293 cells transiently expressing TREK-1. HEK293 cells were cultured in Dulbecco’s Modified Eagle’s Medium supplemented with 10% fetal bovine serum, penicillin, streptomycin, and L-glutamine in 5% CO_2_ at 37° C. They were transfected with 1 μg pIRES2-EGFP containing wild-type TREK-1a using Gene Jammer. To analyze the effects of wild-type (WT) and mutant POPDC2 on TREK-1 currents, 1 μg of pCGI vector containing an empty cassette, or WT or mutant *POPDC2*, were co-transfected. Co-expression of green fluorescent protein (GFP) was achieved by an internal ribosomal entry site cassette in the pIRES2-EGFP vector. Green cells were identified by an epifluorescent microscope and were measured by patch clamp as described below at 48 hours post-transfection.

For the measurement of the cardiac sodium current (I_Na_, encoded by *SCN5A*), a genetically modified HEK-293 cell line stably expressing human Na_V_1.5 channels^17^ was used, cultured under conditions similar to above. The cells were transiently transfected with 1 μg of pCGI containing wild-type *SCN1B* (NM_001037), encoding the β1 subunit of the Na_V_1.5 channel, using Gene Jammer. To analyze the effects of WT and mutant POPDC2 on I_Na_, 1 μg of pCGI vector containing an empty cassette or WT or mutant *POPDC2*, were co-transfected. GFP-positive cells were measured by patch clamp 48 hours post-transfection.

For SDS-PAGE analysis, HEK-293 cells were cultured as described above and transfected with 10 μg of pCGI vector containing an empty cassette or WT or mutant POPDC2 using Lipofectamine 3000 in 6 cm plates and cultured for 48 hours. Protein samples were extracted in sample buffer and protein content evaluated by Lowry method. Forty micrograms of protein treated with 4% β-mercaptoethanol were run on a 10% gradient polyacrylamide gel and then transferred on a nitrocellulose membrane, saturated with Intercept Blocking Buffer PBS (Licor) for one hour at room temperature. The membrane was incubated at 4°C overnight in a blocking solution containing primary antibodies: anti-POPDC2 (Sigma HPA024255, 1:300), GAPDH (SantaCruz sc-20357; 1:2500), GFP (Invitrogen A21311, 1:4000). The next day, the membrane was washed five times with PBS-T (Tween 20, 0.08%) and incubated at room temperature for one hour with the appropriate secondary antibodies in blocking solution (IRDye Licor). The bands corresponding to the proteins of interest were visualized on a Licor Odissey XF imaging system.

### Electrophysiological characterization of POPDC2 variants

#### Data acquisition

I_Na_ and TREK-1 currents were measured with ruptured and amphotericin-perforated patch-clamp technique, respectively, using an Axopatch 200B amplifier (Molecular Devices Corporation, Sunnyvale, CA, USA). Voltage control, data acquisition, and analysis were accomplished using custom software. I_Na_ recordings were low-pass filtered with a cut-off frequency of 5 kHz and digitized at 20 kHz, while this was 2 and 4 kHz, respectively, for TREK-1 current measurements. Series resistance was compensated by ≥80% and potentials were corrected for the calculated liquid junction potential.^18^ Cell membrane capacitance (C_m_) was calculated by dividing the time constant of the decay of the capacitive transient after a −5 mV voltage step from −40 mV by the series resistance. Patch pipettes were pulled from borosilicate glass (Harvard Apparatus, UK) and had resistances of 2.5–3.5 MΩ after filling with the solutions as indicated below.

#### TREK-1 current measurements

TREK-1 currents were recorded at 36±0.2°C. Cells were superfused with solution containing (in mM): NaCl 140, KCl 5.4, CaCl_2_ 1.8, MgCl_2_ 1, glucose 5.5, HEPES 5, pH 7.4 (NaOH). Pipette solution contained (in mM): K-gluc 125, KCl 20, NaCl 5, amphotericin-B 0.88, HEPES 10, pH 7.2 (KOH). TREK-1 currents were measured using 500 ms voltage clamp steps to test potentials ranging from −100 to +50 mV from a holding potential of −80 mV. The TREK-1 current was measured at the end of the voltage clamp step and current densities were calculated by dividing current amplitude by C_m_.

#### I_Na_ measurements

I_Na_ was measured at room temperature using a bath solution containing (in mM): NaCl 20, CsCl 120, CaCl_2_ 1.8, MgCl_2_ 1.0, glucose 5.5, HEPES 5.0, pH 7.4 (CsOH).

Pipettes were filled with solution containing (in mM): NaF 10, CsCl 10, CsF 110, EGTA 11, CaCl_2_ 1.0, MgCl_2_ 1.0, Na_2_ATP 2.0, 10 HEPES, pH 7.2 (CsOH). The I_Na_ density and voltage dependence of activation were determined by 50 ms depolarizing pulses to test potentials ranging from −80 to +40 mV from a holding potential of −120 mV. Voltage-dependent inactivation was obtained by measuring the peak currents during a 50 ms test step to −20 mV, which followed a 500 ms prepulse to membrane potentials between −140 and 0 mV to allow inactivation. The holding potential was −120 mV. All voltage clamp steps were applied with a 5 s cycle length. Peak I_Na_ was defined as the difference between peak and steady-state current. Current density was calculated by dividing the measured currents by C_m_. To determine the activation characteristics of I_Na_, current-voltage curves were corrected for differences in driving force and normalized to maximum peak current (I_max_). Steady-state activation and inactivation curves were fitted using the Boltzmann equation I/I_max_=A/{1.0+exp[(V_1/2_-V)/k]} to determine the membrane potential for half-maximal (in)activation (V_1/2_) and the slope factor k.

#### Statistical analysis

Data are presented as mean ± standard error of the mean (SEM). Statistical analysis was carried out with SigmaStat 3.5 software. Normality and equal variance assumptions were tested with the Kolmogorov-Smirnov and the Levene median test, respectively. Groups were compared with one-way ANOVA. *P*<0.05 defines statistical significance.

### Computer simulations

The spontaneous electrical activity of a single human sinus nodal pacemaker cell was simulated using the comprehensive mathematical model developed by Fabbri et al.^19^ For simulations of a single human atrial cell, we used the model by Maleckar et al.^20^ The CellML code of both models, as available from the CellML Model Repository^21^ at https://www.cellml.org/, was edited and run in version 0.9.31.1409 of the Windows based Cellular Open Resource (COR) environment.^22^ TREK-1 currents are not included in both original models. To study whether the homozygous loss-of-function variants in *POPDC2* contribute to bradycardia via TREK-1 current changes, we fitted our experimental data of the TREK-1 current-voltage relationship (**Figure 3A**) and implemented the thus obtained TREK-1 current in both models as a control, over a range of TREK-1 current densities. Subsequently, the TREK-1 density was reduced to 59% of the TREK-1 + POPDC2*-*WT current according to the effects induced by the *POPDC2* variants (**Figure 3A**) to assess the functional effects of the variants. All simulations were run for a period of 200 s, which appeared a sufficiently long time to reach steady-state behavior. The analyzed data are from the final 10 seconds of the 200 s period.

### Association of heterozygous *POPDC2* variants and burden analyses for *POPDC2* with clinical phenotypes and heart rate in population biobanks

#### Biobank description and phenotyping

Samples were included from four large population biobanks (total n=1,089,031) with genetic data, namely deCODE genetics in Iceland (n=173,025), UK Biobank (n=428,503), Copenhagen Hospital Biobank and the Danish Blood Donor study in Denmark (n=487,356) and Intermountain in Utah, USA (n=138,006). Disease status was obtained from electronic health records and ascertained using the following International Classification of Diseases 10th (ICD10) revision codes: Atrioventricular block (I44.1 and I44.2), Bradycardia (R00.1), Cardiac arrest (I46), Hypertrophic cardiomyopathy (I42.1 and I42.2), Muscular dystrophy (G71.0), Myocarditis (I40, I41 and I51.4), Sinus node dysfunction (I49.5). Pacemaker implantation was defined based on procedure codes (deCODE: Nomesco Classification of Surgical Procedures (NCSP) codes FPE/FPSE and FPF/FPSF, Copenhagen Hospital Biobank: NCSP codes FPE/FPSE and FPF/FPSF, UK Biobank: OPCS code K60). Heart rate was available only in the UK Biobank and deCODE. In the UK Biobank, heart rate was obtained during blood pressure measurement at assessment. Both measurements were taken twice and multiple measurements for one individual were averaged. In deCODE, heart rate measurements were sourced from ECGs from Landspitali University Hospital in Iceland between 1998 and 2015. Mean values from sinus rhythm ECGs were obtained for each individual, which were subsequently standardized and adjusted for age and sex. Details on each biobank and DNA genotyping and sequencing methods for each biobank can be found in the **Supplemental Methods**.

#### Gene burden models

We defined different models to group together various types of variants:

A. LOF: only loss-of-function (LOF) variants according to Variant Effect Predictor (VEP)^23^
B. LOFTEE: high-confidence LOF variants according to LOFTEE^24^
C. LOFCADD: LOF and missense (MIS) when predicted deleterious with CADD phred score ≥ 25^25^
D. LOF1MISID: LOF and MIS when predicted deleterious by at least one of the following prediction methods: MetaSVM, MetaLR^26^ or CADD phred score ≥ 25.^25^
E. all models, we used MAF < 2% to select variants for analyses.

#### Association analyses

For case-control analyses, we used logistic regression under an additive model to test for association between carrying a loss-of-function variant in *POPDC2* (LOF, MIS according to each model A-D) and phenotypes, in which disease status was the dependent variable and genotype counts as the independent variable. For the analyses, we used software developed at deCODE genetics.^27^ For testing association with heart rate, measurements were inverse-normal transformed and analyzed using a linear mixed model implemented in BOLT-LMM.^28^ Meta-analysis was performed on the summary results from IS, UK, DK and US when available, using a fixed effects inverse variance weighted method.^29^

## RESULTS

### *POPDC2* variants cause sinus node disease and AV conduction defects with variable hypertrophic cardiomyopathy in multiple families

To uncover the genetic cause of sinus node disease, AV conduction defects and HCM in a child (age at presentation 11-15 years, Family A) we performed whole-exome sequencing in the proband, both parents and two of his unaffected siblings. His parents are first cousins (**Figure 1**, individual II-3 in the pedigree, **Table 1**, **Supplemental Figure 1**).

**Figure 1.**
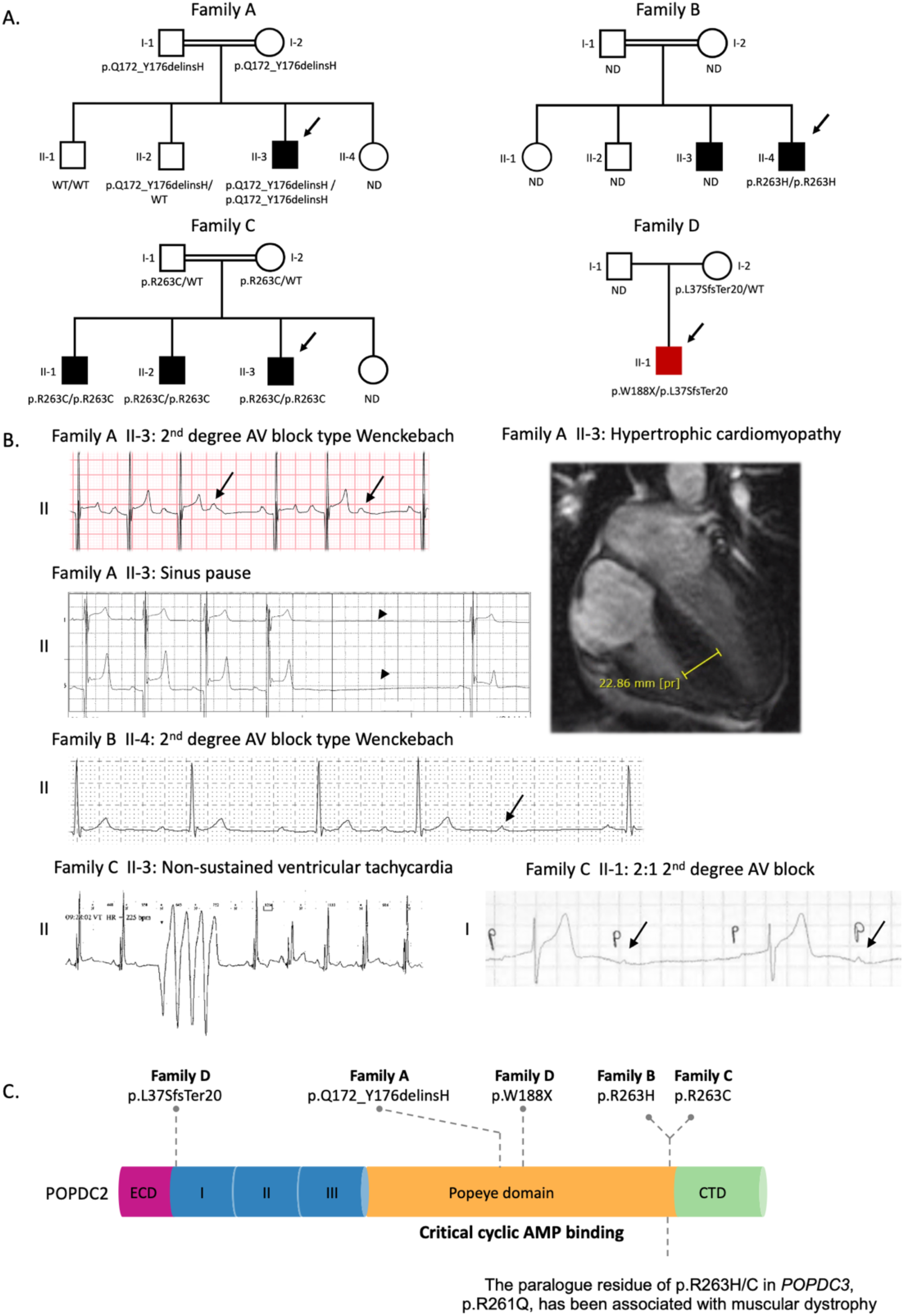
Bi-allelic variants in *POPDC2* cause a recessive syndrome with sinus node disease and atrioventricular conduction defects with variable hypertrophic cardiomyopathy. **(A)** Pedigrees of Families A-D. Closed symbols indicate affected individuals. Males are indicated by squares and females by circles. A double line indicates a consanguineous relationship. The arrows points to the probands. **(B)** Selection of electrocardiographic (ECG) abnormalities: (1) patient II-3 from Family A (2^nd^ degree AV block type Wenckebach and sinus pause [indicated by an arrow head,] see **Supplemental** Figure 1 for longer Holter registration), (2) patient II-4 from Family B (2^nd^ degree AV block type Wenckebach), patient II-3 (non-sustained ventricular tachycardia) and patient II-1 (2:1 2^nd^ degree AV block) from Family C. Arrows point to a non-conducted P wave. **Upper right panel:** Cardiac MRI at the age of 11-15 years showing marked hypertrophy of the interventricular septum (23 mm, Z-score^48^: 16.43; height, 160 cm; weight, 49 kg) in the proband of Family A. **(C)** POPDC2 protein domain structure and location of variants found homozygously in patients. **Abbreviations:** AV, atrioventricular; CTD, carboxy-terminal domain; ECD, extracellular domain; ND, genotype not determined; Regions I/II/II, transmembrane region 1-3; WT, wild-type.

**Table 1.** Clinical characteristics of patients with biallelic *POPDC2* variants.

|  | Family A | Family B | Family C |  |  | Family D |
| --- | --- | --- | --- | --- | --- | --- |
|  | II-1 | II-1 | II-1 | II:2 | II:3 | II-1 |
| ***POPDC2* variant** | p.Q172_Y176delinsH | p.R263H | p.R263C |  |  | p.W188X/p.L37SfsTer20 |
| **Effect on TREK1 current** | Loss of function | Loss of function | Not tested |  |  | Not tested |
| **Consanguinity** | Yes | Yes | No |  |  | No |
| **Sex** | M | M | M | M | M | M |
| **Status** | Alive | Alive | Alive | Alive | Alive | Alive |
| **Age at presentation (years)** | 11-15 | 21-25 | 21-25 | 21-25 | 16-20 | 11-15 |
| **Clinical symptoms at presentation** | Palpitations | Palpitations | None | None | Cardiac arrest | Chest pain |
| **Cardiac arrhythmia and ECG abnormalities** |  |  |  |  |  |  |
| **Cardiac arrest** | No |  | No | No | Arrest following bradycardia | Arrest following bradycardia |
| **Minimum heart rate (BPM)** | 33 | 30 | 33 | NA | NA | 67 |
| **Sinus node disease** | Sinus pauses (up to 3s) | Sinus bradycardia | No | SA-block with sinus pauses (up to 5s) | SA-block Sinus arrest | No |
| **AV-conduction disease** | 2nd degree AV-block (Type 1) | PQ time of 200ms<br>2nd degree AV-block (Type 1) | 1st degree AV-block<br>2nd degree AV-block (Type 1 and 2:1) | 1st degree AV-block | 1st degree AV-block<br>2nd degree AV-block (Type 1) | 1st degree AV-block |
| **Other ECG abnormalities** | No | No | No | Right bundle branch block | No | Diffuse ST-segment elevation |
| **Atrial arrhythmia** | Atrial fibrillation and flutter | No | Atrial fibrillation and flutter | No | Episodes of high atrial rate | No |
| **Ventricular arrhythmia** | Monomorphic NSVT, PVCs | No | Monomorphic NSVT | No | Monomorphic NSVT | VT |
| **Exercise ECG** | 2nd degree AV-block (Type 1) in recovery phase | Normal | NA | NA | NA | NA |
| **Structural cardiac and extra-cardiac abnormalities** |  |  |  |  |  |  |
| **Hypertrophic cardiomyopathy** | Yes | Yes | No | No | No | No |
| **Cardiac MRI findings** | Septal hypertrophy (23 mm) | Septal hypertrophy (16mm) | NA | NA | Possible myocarditis | Significant LV fibrosis and inflammation |
| **TTE (septal thickness)** | Septal hypertrophy (23 mm) | Septal hypertrophy (11mm) | Normal | Normal | Normal | At presentation: Severe biventricular dysfunction. Follow-up: Moderate LV systolic dysfunction without dilatation or hypertrophy, and normal RV size and function. |
| **Myocarditis** | No | No | Np | No | Possible myocarditis | Clinically (not on cardiac histology) |
| **Muscular dystrophy** | No | No | No | No | No | No |
| **Cardiac biopsy** | NA | NA | Normal | NA | Fatty tissue and fibrosis | Non-specific fibrosis |
| **Treatment** |  |  |  |  |  |  |
| **Pacemaker implantation (age, years)** | Yes (11-15) | No | Yes (21-25) | Yes (21-25) | Yes (16-20) | Yes (11-15), temporary |
| **ICD implantation (age, years)** | Yes (11-15) | No | No | No | Yes (31-35) | No |
| **Appropriate ICD shocks** | No | NA | NA | NA | No | NA |

**Abbreviations:** AV, atrioventricular, BPM, beats per minute; ICD, implantable cardiac defibrillator; MRI, magnetic resonance imaging; NA, not available, NSVT, non-sustained ventricular tachycardia; PVC, premature ventricular contractions; SA, sinoatrial; TTE, transthoracic echocardiogram; VT, ventricular tachycardia

No likely causal variant was found in genes previously associated with Mendelian cardiomyopathies or arrhythmia syndromes (either recessive or dominant). In line with the expected recessive inheritance pattern, we identified a rare segregating homozygous variant, p.Q172_Y176delinsH (GenBank accession number, NM_022135), in the Popeye domain-containing protein 2 *(POPDC2),* as the most likely variant consistent with a recessive mode of inheritance and parental consanguinity (see **Supplemental Table 3** for overview of the two segregating coding region variants identified in the homozygous state). Of note, the only other homozygous segregating variant has been found 15 times in gnomAD and was therefore found unlikely to cause the disease in this patient. No *de novo* variants were found in the index patient. The cardiac arrhythmia phenotype in the patient is consistent with studies in mice that showed that loss of *Popdc2* resulted in sinus pauses and bradycardia^5^, and with findings made in zebrafish where morpholino knockdown of *popdc2* resulted in AV block.^6^ We therefore considered the p.Q172_Y176delinsH homozygous variant in *POPDC2* as an excellent candidate for this novel cardiac disorder. We then searched for additional patients carrying biallelic variants in *POPDC2* by screening this gene in 78 patients that presented with a similar phenotype as Family A (i.e. CCD with HCM) and a search using GeneMatcher^8^ and Decipher^30^. In total, we identified 6 patients from 4 unrelated families of different ancestries (patients 3-5 are siblings, **Figure 1A-B**, **Table 1**) carrying either homozygous or compound heterozygous rare *POPDC2* variants. All 6 patients were diagnosed with cardiac conduction defects with or without HCM.

In summary (see **Table 1**, **Figure 1B**), (1) AV conduction disease was present in all 6 patients, mainly consisting of 1^st^ degree AV block or 2^nd^ degree AV block type 1 (Wenkebach), (2) sinus node disease presenting as sinus bradycardia and sinus pauses was present in 4/6 patients, (3) cardiac arrest accompanied by sinus bradycardia or asystole was seen in 2/6 patients, (4) atrial arrhythmia (i.e. atrial flutter or fibrillation) was detected in 3/6 patients, (5) non-sustained ventricular tachycardia occurred in 4/6 patients and (6) HCM was diagnosed in 2/6 patients (probands in Families A and B; with no other genetic variant causing hypertrophic cardiomyopathy). A pacemaker was implanted in 5/6 patients with the age at implantation ranging from 15-23 years. In 2/6 patients an implantable cardioverter-defibrillator (ICD) was implanted to prevent lethal ventricular arrhythmia, but no appropriate ICD shocks occurred. Unlike the recessive syndrome associated with the other POPDC genes (i.e. *POPDC1* and *POPDC3-related limb-girdle muscular dystrophy)*, none of the patients showed signs of muscular dystrophy. Of note, patient 6 (Family D) presented with acute myocarditis for which he was admitted to the intensive care unit. Detailed phenotypic descriptions of all patients can be found in **Supplemental Note 1.**

We noted an autosomal-recessive mode of inheritance in all the families and found that variants in *POPDC2* were inherited from each of the unaffected parents, but unknown for patients 2 and 6 (from Family B and D, **Figure 1A**) for which DNA of (one of) the parents was not available. In total, we report 5 *POPDC2* variants, of which 2 are missense variants (p.R263H, p.R263C), 1 in-frame insertion deletion (p.Q172_Y176delinsH) and 2 are expected to result in protein truncation (p.W188X, p.L37SfsTer20, **Supplemental Table 2**). The minor allele frequency (MAF) of *POPDC2* variants identified in patients ranged from 1.4×10^-5^ to 1.3×10^-4^ in the gnomAD v4.0 (accessed March 2024, **Supplemental Table 2**). Furthermore, none of the variants were found homozygous in 730,947 exomes and 76,215 genomes from gnomAD, suggesting that homozygosity for these variants is not well tolerated. In line with this, all variants are predicted damaging by multiple *in silico* prediction tools (**Supplemental Table 2**).^11^ The variant identified in Family A results in an in-frame deletion of 5 highly conserved amino acid residues at positions 172-176 and the insertion of a histidine at position 172 of the POPDC2 protein (p.Q172_Y176delinsH, **Figure 1C, Supplemental Table 2**). The variant identified in Family B replaces the highly conserved arginine residue at position 263 by a histidine (p.R263H, **Figure 1C**). Intriguingly, we identified p.R263C in family C affecting the same residue as the variant found in Family B but mutating Arg to Cys instead of Arg to His. Of note, the paralogous residue of p.R263H/p.R263C in *POPDC3*, p.R261Q, has in the homozygous state been recently associated with muscular dystrophy without cardiac arrhythmia.^31^ In Family C, the three affected siblings homozygous for p.R263C, inherited the variant from the heterozygous and clinically unaffected parents. We performed a look-up of the p.R263C variant in recent whole-genome sequencing dataset^32^ consisting of young patients (n=226) receiving a pacemaker because of AV block and none of the patients was homozygous for p.R263C nor were there any patients with bi-allelic variants in *POPDC2* (**Supplemental Note 2**). In Family D we identified p.W188X and p.L37SfsTer20 in compound heterozygosity that are expected to result in loss of function of the gene due to premature truncation of the protein and nonsense -mediated decay. The cardiac arrest accompanied by sinus bradycardia and 1^st^ degree AV block he displayed fit the conduction disease seen in the other *POPDC2* patients. However, although the cardiac biopsy was negative for myocarditis, a potential causal role of myocarditis cannot be ruled out. Nevertheless, as loss of *Popdc2* in mice results in sinus pauses and bradycardia^5^ and morpholino knockdown of *popdc2* resulted in AV block, we considered bi-allelic truncating variants as strong evidence of pathogenicity. Moreover, gnomAD contains 70 predicted loss-of-function *POPDC2* variants; none of them occurs in the homozygous state, suggesting that *POPDC2* is intolerant to homozygous loss-of-function variants. This is further supported by the previous observation that homozygous knock-in mice of the p.W188X variant (found in compound heterozygosity in Family D) displayed stress-induced sinus bradycardia and pauses, consistent with the phenotype in patient 6 and the other five patients we report on. There was no individual among 125,748 exomes and 15,708 genomes in Genome Aggregation Database v2.1.1 (gnomAD) that carried both variants found in compound heterozygosity (p.W188X and p.L37SfsTer20) in the patient from Family D. In aggregate, these data suggest a causal role for these variants in this patient, with myocarditis possibly playing a precipitating role in the presentation of the cardiac conduction defects.

### Homology model for POPDC2

To explore the structural and functional consequences of the homozygous and compound heterozygous *POPDC2* variants, we generated a predicted structural model of POPDC2, as no experimentally determined structures are currently available. SWISS-MODEL^9^ and AlphaFold Multimer^10^ were used since these protein structure prediction programs could generate dimeric models of POPDC2, and dimerization has been shown to be critical for the activity of POPDC1^26^. As a complementary approach, AlphaMissense^11^ was used to predict relative pathogenicity scores of the identified single amino acid substitutions and deleted regions.

AlphaFold Multimer generated a high confidence dimer **(Supplemental Figure 2A)** for full-length POPDC2 with a dimeric transmembrane domain composed of 6 transmembrane α-helices (3 α-helices from each subunit) and two cAMP-binding Popeye domains that also contained an extensive dimer interface **(Figure 2A)**. In the AlphaFold model, Arg263 (mutated in Families B and C) was pointing into the cAMP-binding pocket of the Popeye domain from the adjacent subunit. SWISS-MODEL was the only program able to model a cAMP-bound state of the Popeye domain **(Supplemental Figure 2B)**. To generate a model of the cAMP bound state of full-length POPDC2, we merged the AlphaFold Multimer and SWISS-MODEL predictions by swapping the Popeye domains (see methods). This created a merged model of POPDC2 that included the transmembrane domain and the Popeye domains bound to cAMP **(Figure 2A)**.

**Figure 2.**
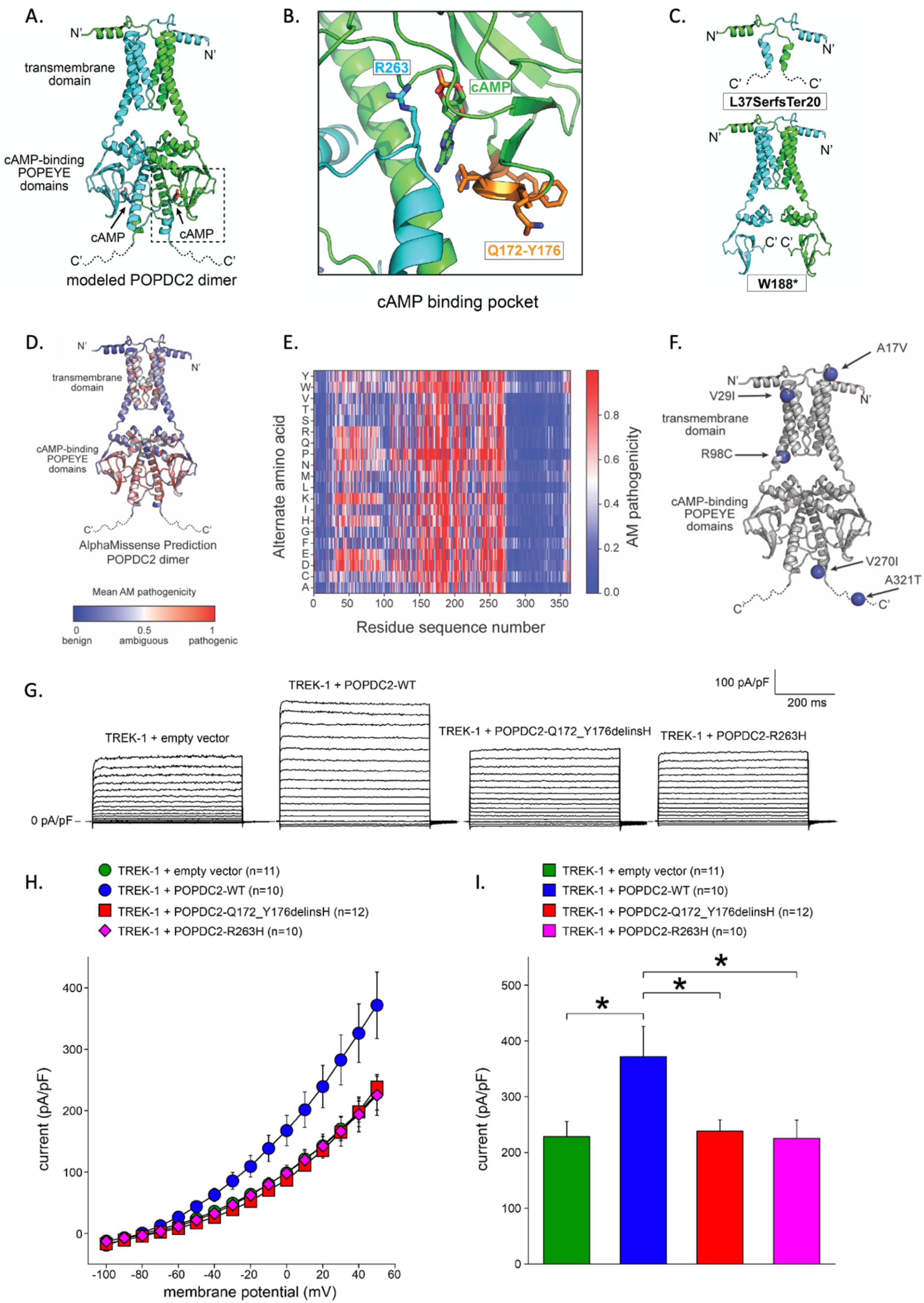
**Functional characterization of the *POPDC2* variants** (A) Structural model of POPDC2 bound to cAMP generated using AlphaFold2 Multimer and SWISS-MODEL. Dimer subunits are shown in green and cyan; cAMP molecules, green and cyan sticks. N’ and C’ indicate the N- and C-termini. The positions of the transmembrane and Popeye domains are indicated by the labels. The intrinsically disordered C-terminus (residues 275-364) are shown as dotted lines. (B) Zoom-in of the predicted cAMP binding pocket of POPDC2. Dimer subunits are shown in green and cyan; cAMP, green sticks; residues p.Q172-Y176, orange; p.R263, cyan sticks. (C) Structural Models of the POPDC2 variants p.L37SfsTer20 and p.W188X, which would both generate truncated proteins that would lack the ability to bind cAMP. (D) Homology model of POPDC2 by AlphaFold Multimer color coded by the average pathogenicity score for each residue as predicted by AlphaMissense. (E) Heatmap of predicted effects of amino acid substitutions on POPDC2. AlphaMissense (AM) scores range from zero to one, with higher scores corresponding to increased pathogenicity. (F) Homology model of POPDC2 by AlphaFold Multimer color coded by the average pathogenicity score for each residue as predicted by AlphaMissense. The position of non-disease associated variants found in general population are shown as blue spheres, indicating a predicted AlphaMissense pathogenicity scores <0.1. (G) Typical examples of TREK-1 currents upon 500 ms voltage clamp steps to membrane potentials ranging from −100 to +50 mV from a holding potential of −80 mV in absence or presence of wild-type (WT) and mutant POPDC2. (H) Average current-voltage relationships of TREK-1 currents in absence or presence of WT and mutant POPDC2. (I) TREK-1 current amplitude at +50 mV in absence or presence of WT and mutant POPDC2. \**P*<0.05 with one-way ANOVA.

Based on the structural model, all variants (p.Q172-Y176delinsH, p.R263H, p.R263C and p.W188X/p.L37SfsTer20) are predicted to diminish the ability of POPDC2 to bind cAMP. The first variant (p.Q172-Y176delinsH, Family A) replaces five residues with a single histidine residue. The deleted residues are predicted to form one beta strand that is part of a three stranded beta sheet at the base of the cAMP binding pocket **(Figure 2B)**. Replacement of the 5 residues with a single histidine residue would significantly alter the structural integrity of the cAMP-binding Popeye domain and directly affect the ability of POPDC2 to bind cAMP.^26^ The second and third variants, p.R263H (found in Family B) and p.R263C (found in Family C), are also both predicted to interfere with proper cAMP binding, as the positively charged Arg residue is predicted to be near the negatively charged cyclic phosphate group of cAMP **(Figure 2B)**. Mutation of the Arg to either His or Cys would eliminate a key interaction predicted to stabilize cAMP binding and thus reduce or eliminate the ability of POPDC2 to bind cAMP. In support of R263 being involved in a specific interaction to stabilize cAMP binding, mutation of R263 to any other residue is predicted to be pathogenic by AlphaMissense, with pathogenicity scores of 0.82 and 0.79 for the R263H and R263C variants, respectively.

The compound heterozygous variants are also predicted to eliminate cAMP binding by POPDC2 **(Figure 2C)**. The L37SerfsTer20 variant is a truncated protein that completely lacks the cAMP-binding Popeye domain and would also generate an incomplete transmembrane domain. The p.W188X variant would retain the dimeric transmembrane domain, but truncate the Popeye domain, thus leaving an incomplete domain incapable of binding to cAMP **(Figure 2C)**.

Notably, residues with high pathogenicity scores (>0.5) all cluster in the regions of the POPDC2 structure that are involved in either dimerization or cAMP binding **(Figure 2D-E)**, which emphasizes the importance of cAMP binding and dimerization for POPDC2 function. Consistently, other variants within POPDC2, found homozygously in gnomAD^12^, and without a clear disease association, (p.A17V, p.V29I, p.R98C, p.V270I, and p.A321T) are all positioned away from the cAMP binding pocket and are predicted to benign by AlphaMissense with low pathogenicity scores <0.1 (**Figure 2E-F**). Taken together, the disease-associated variants reported here are primarily located in regions of the POPDC2 structure that are critical for protein function.

### *POPDC2* variants fail to increase TREK-1 current density

We then aimed to functionally characterize the *POPDC2* variants. TREK-1 is a recognized interacting protein of POPDC2^5^ and co-expression of POPDC2 and TREK-1 has been shown to increase TREK-1 current in comparison to expression of TREK-1 alone.^5^ Furthermore, cardiac-specific TREK-1 deficient mice display a sinus node phenotype characterized by bradycardia with frequent episodes of sinus pauses, partially resembling the phenotype in the patients with homozygous *POPDC2* variants presented here.^33^ We therefore hypothesized that the effect of the p.Q172_Y176delinsH (Family A) and p.R263H (Family B) variants in *POPDC2* is mediated through modulation of the TREK-1 current. We did not test the variants found in Family C (p.R263C) and Family D (p.W188X/p.L37SfsTer20) as they affect the same residue as the variant in Family B (p.R263H) or are expected to result in premature truncation of the protein, respectively. We co-transfected HEK-293 cells with WT and mutant POPDC2 with TREK-1 containing plasmids. As expected^5^, co-expression of WT POPDC2 with TREK-1 increased TREK-1 current density (**Figure 2G-I**). However, when we co-expressed TREK-1 with a p.Q172_Y176delinsH or p.R263H POPDC2-containing plasmid, no increase in TREK-1 current density was observed, comparable to the co-expression of TREK-1 with an empty vector (**Figure 2G-I**). We then tested the effect of the *POPDC2* variants on sodium current (I_Na_) as it was recently hypothesized that POPDC2 may interact with Na_V_1.5.^34^ However, expression of WT, p.Q172_Y176delinsH or p.R263H POPDC2 in a HEK-293 cell line stably expressing human Na_V_1.5 channels (encoded by *SCN5A*) showed neither an effect on I_Na_ density nor on I_Na_ gating properties by WT or mutant POPDC2 (**Supplemental Figure 3**). Western blot analysis showed levels of expression of the mutant proteins that were comparable to the levels seen for WT POPDC2 (**Supplemental Figure 4**).

### *In silico* modeling of the decrease in TREK-1 current induced by the POPDC2 variants on human sinus node and atrial cells

To evaluate whether the observed decrease in TREK-1 current density, associated with the homozygous *POPDC2* variants, is responsible for the bradycardia observed in patients, we conducted computer simulations using comprehensive mathematical models of both a human sinus nodal pacemaker cell^19^ and a human atrial cell.^20^ As shown in **Figure 3A** (blue line), the experimental data on the voltage dependence of the TREK-1 + POPDC2-WT current could be well-fitted (r^2^>0.99) with the relationship I_TREK-1_ = −117.1 + 284.24 × exp(V_m_/90.52), in which I_TREK-1_ and V_m_ denote TREK-1 current density (in pA/pF) and membrane potential (in mV), respectively. The mutant data could be well-fitted by scaling down I_TREK-1_ to 59% of the TREK-1 + POPDC2-WT current over the entire volage range (**Figure 3A**, red line). We started our simulations with incorporating I_TREK-1_, which is not present in the original model cell, into the human sinus node model cell. Because data on the density of I_TREK-1_ in human sinus node cells are lacking, we set its density to 1.2 pA/pF (at +30 mV), as observed in mouse sinus node cells by Unudurthi et al.^33^ This, however, resulted in cessation of pacemaker activity in the human sinus node model cell. We then repeated our simulations with a 10 times lower density of I_TREK-1_. With this density the cycle length of the simulated action potential amounted to 1160 ms (**Figure 3B**, top panel, blue solid line), as compared to 813 ms in the original model (**Figure 3B**, top panel, grey dotted line). Cycle length was reduced by 16% from 1160 to 973 ms upon reduction of I_TREK-1_ density to 59%, thus simulating the effect of the variants in *POPDC2* (**Figure 3B**, top panel, red solid line, and **Figure 3C**, vertical arrow). The decrease in cycle length was mainly due to an increase in diastolic depolarization rate (**Figure 3B**, top panel) as a result of the decrease in the small but effective I_TREK-1_ during this phase of the action potential (**Figure 3B**, bottom panel). Qualitatively similar results were obtained with other I_TREK-1_ densities. The observed effects on cycle length are summarized in **Figure 3C**.

**Figure 3.**
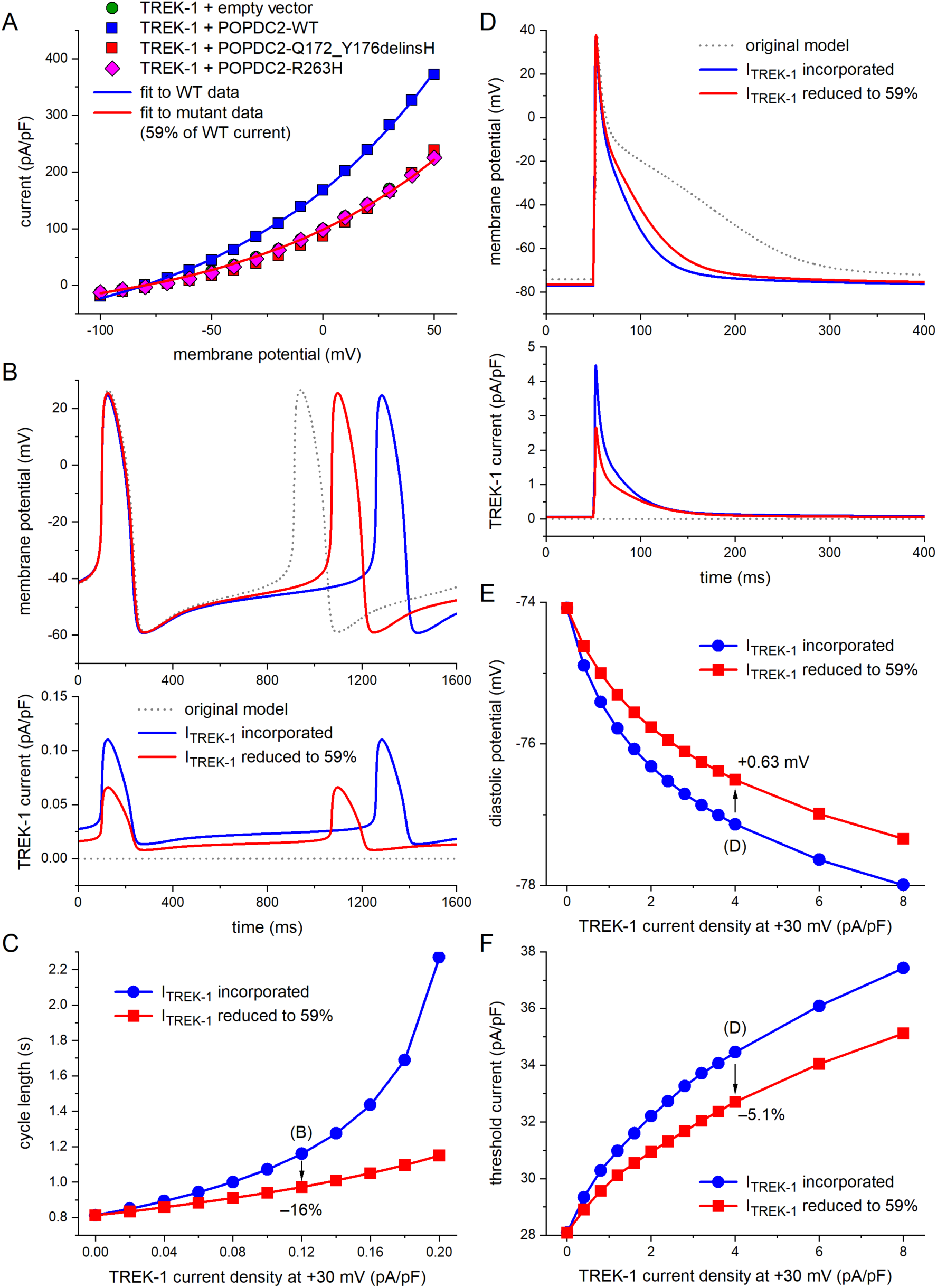
**Functional effects of the *POPDC2* variants from *in silico* modeling** (A) Fits to the experimental data on TREK-1 currents in absence or presence of wild-type (WT) and mutant POPDC2. The fit to the mutant data was obtained by scaling the fit to the WT data by a factor of 0.59. (B) Membrane potential (top) and associated TREK-1 current (bottom) of a single human sinus nodal pacemaker cell as simulated using the comprehensive mathematical model developed by Fabbri et al.^19^ I_TREK-1_ was introduced into the original model cell using the fits of panel A. I_TREK-1_ magnitude was set to 0.12 pA/pF at a membrane potential of +30 mV. (C) Cycle length of the simulated single human sinus nodal pacemaker cell as a function of I_TREK-1_ magnitude. (D) Membrane potential (top) and associated TREK-1 current (bottom) of a single human atrial cell as simulated using the comprehensive mathematical model developed by Maleckar et al.^20^ I_TREK-1_ was introduced into the original model cell using the fits of panel A. I_TREK-1_ magnitude was set to 4.0 pA/pF at a membrane potential of +30 mV. Action potentials were elicited at a rate of 1 Hz with a 1-ms, 20% suprathreshold stimulus current. (E) Diastolic potential of the simulated single human atrial cell as a function of I_TREK-1_ magnitude. (F) Threshold stimulus current of the simulated single human atrial cell as a function of I_TREK-1_ magnitude.

One may argue that the *POPDC2* variants-induced decrease in I_TREK-1_ affects pacemaker activity by changing the excitability of the atrial tissue surrounding the sinus node. This was assessed in simulations of a human atrial cardiomyocyte after incorporating I_TREK-1_, which was not represented in the original model cell.^20^ **Figure 3D** shows the results obtained with an I_TREK-1_ density of 4.0 pA/pF (at +30 mV). Simulating the effect of the variants in *POPDC2* by lowering the density of I_TREK-1_ to 59% led to a 0.63 mV depolarization of diastolic potential, a 5.1% decrease in threshold current (**Figure 3**, **E**-**F**, vertical arrows), and a 23 ms increase in action potential duration (APD) at 90% repolarization. The relatively small effects on diastolic potential and threshold current were obtained with a probably overestimated I_TREK-1_ density, given the large effect of I_TREK-1_ on the APD of the original model (**Figure 3D**, top panel). Smaller, but qualitatively similar effects on diastolic potential and threshold current were obtained with lower I_TREK-1_ densities (**Figure 3E**-**F**), associated with a less prominent effect of I_TREK-1_ on the APD of the original model. Overall, despite the observation of bradycardia in mice lacking TREK-1, findings from our simulation studies do not provide an explanation for the clinically observed sinus bradycardia of the *POPDC2* variants based on a decrease in I_TREK-1_ per se.

### p.Q172-Y176delinsH results in a significant reduction of both POPDC1 and POPDC2 expression in skeletal muscle

Patient II-3 from Family A underwent muscle biopsy of the m. vastus lateralis. Histological analysis disclosed mild fiber size variability and a slight increase of connective tissue. Neither nuclear centralizations nor fiber splittings were observed. No necrotic or degenerated fibers were detected (**Figure 4A and B**). Non-specific myopathic features and increased connective tissue were previously observed in muscle samples of *POPDC1* patients^35–37^, while patients with *POPDC3* variants display typical features of muscle dystrophies, although the severity of the histopathological findings differed among patients.^31^

**Figure 4.**
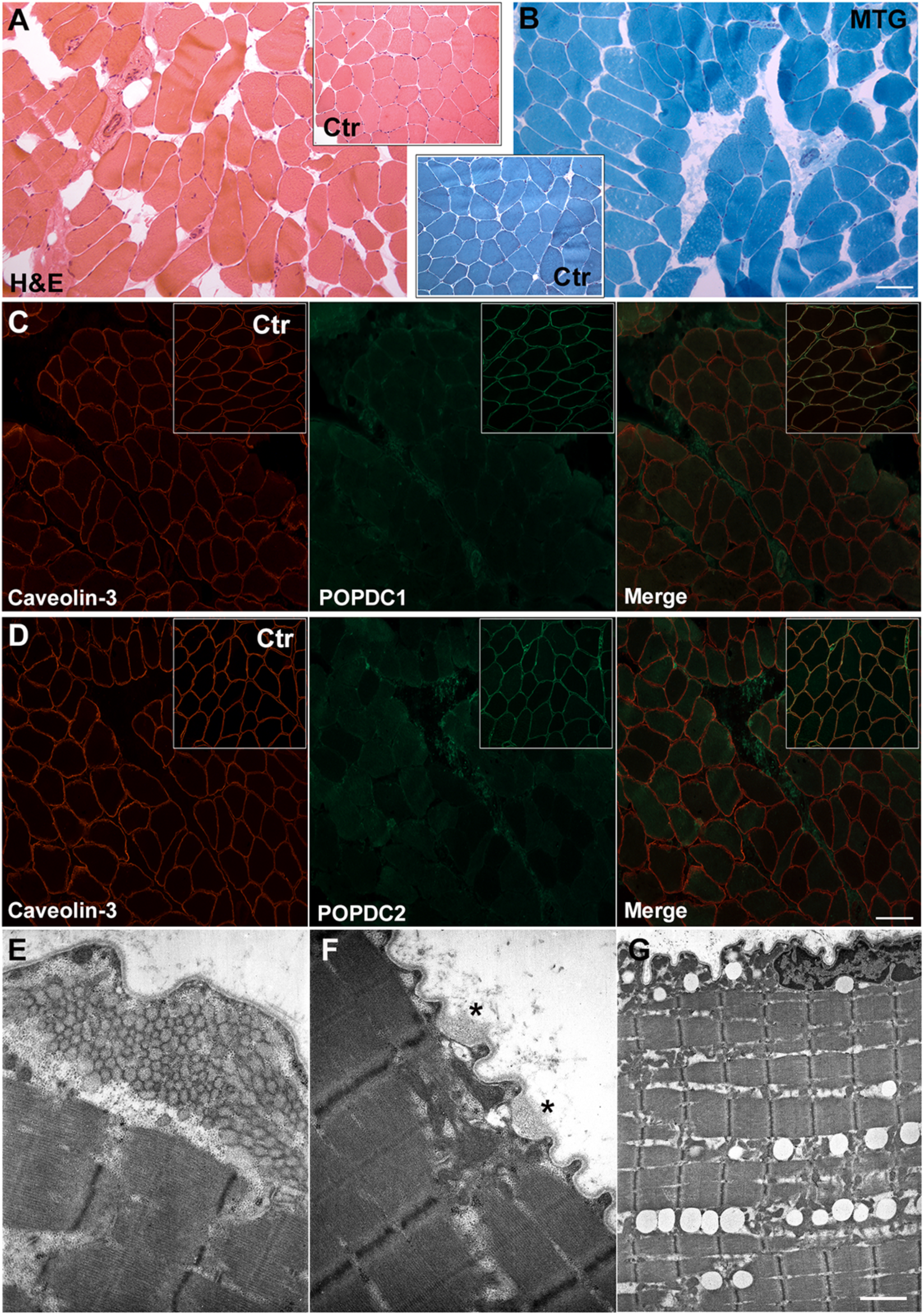
Evaluation of muscle biopsy from the proband in Family A. **(A)** Hematoxylin and eosin (H&E) stain and **(B)** Modified Gomori Trichrome (MGT) staining of patient and, in the inset, control muscle. Scale bar 50 μm. **(C)** Immunofluorescent staining for caveolin-3 (red), POPDC1 (green) and merge for patient. The inset shows the corresponding immunofluorescence staining for control. Scale bar 50 μm. **(D)** Immunofluorescent staining for caveolin-3 (red), POPDC2 (green) and merge for patient. The corresponding control staining is shown in the inset. Scale bar 50 μm. **(E-G)** Ultrastructural findings. **(E)** Tubular aggregates in subsarcolemmal region. **(F)** Sarcolemma alteration (asterisks). **(G)** Increase in lipid droplets. Scale bar E, F: 0.84 μm, G: 3.33 μm.

In healthy skeletal muscle fibers, POPDC1 and POPDC2 were robustly expressed in the sarcolemma. Conversely, in the patient’s muscle, immunofluorescence staining showed a significant reduction of POPDC1 (**Figure 4C**) and POPDC2 (**Figure 4D**) levels. On the other hand, the immunofluorescent signal for caveolin-3, which also stains muscle membranes, showed normal distribution and intensity in patient’s muscle sample compared to control. The severe reduction of POPDC1 and POPDC2 levels was also documented by SDS-PAGE analysis of muscle protein lysates (**Supplemental Figure 5**). These findings suggest that the *POPDC2* c.516_528delGTTTCTGCACTA variant affects the stability of the POPDC2 protein, hampering its membrane localization and leading to the secondary reduction of POPDC1.

Indeed, the stability and/or membrane trafficking of the POPDC1–POPDC2 complex have been found to be impaired by genetic variants in each of the two proteins.^36,37^

Ultrastructural examination detected the presence of tubular aggregates in few muscle fibers (**Figure 4E**). Alterations in the structure of the sarcolemma, characterized by small microvilli-like projections, together with subsarcolemmal vacuoles were also observed. Basal lamina appeared unstructured and enlarged in some fibers (**Figure 4F**, asterisks). We also noted increased level of lipids, which in some cases were arranged to form rows of droplets (**Figure 4G**). Heterogeneous transmission electron microscopy findings were previously observed in muscle samples from patients harboring variants in POPDC1^35,36^, but not in *POPDC3*-mutated patients.^31^

### POPDC2 expression in human hearts

To explore the anatomical distribution of POPDC1-3 expression (POPDC1’s official gene name is BVES), we analyzed a previously published Visium Spatial and single-nucleus transcriptomic dataset.^14^ For the Visium Spatial dataset, annotation of histological tissue sections of the AV node and the sinus node was used and expression of *POPDC1-3* was explored across those structures (**Figure 5A**). POPDC2 showed higher expression than POPDC1/BVES and POPDC3. Cardiomyocyte rich structures showed higher POPDC2 expression compared to cardiomyocyte poor regions (e.g. cardiac skeleton or fat) (**Figure 5B**). Swan et al. described the necessity of POPDC1-2 co-expression for their trafficking to the cell membrane which is required for their proper function.^37^ Variants in POPDC1/BVES–POPDC2 have also been found to affect the stability and/or membrane trafficking of the complex^36,37^ as corroborated here in the muscle biopsy of the patient from Family A (**Figure 4**). We therefore interrogated the co-expression pattern of POPDC1/BVES and POPDC2. The most prominent area where POPDC1/BVES and 2 expression was seen co-localising in the same Visium spots was the AV node (**Figure 5C**). The sinus node expressed POPDC2, but expression of POPDC1 was sparse (**Figure 5B**).

**Figure 5.**
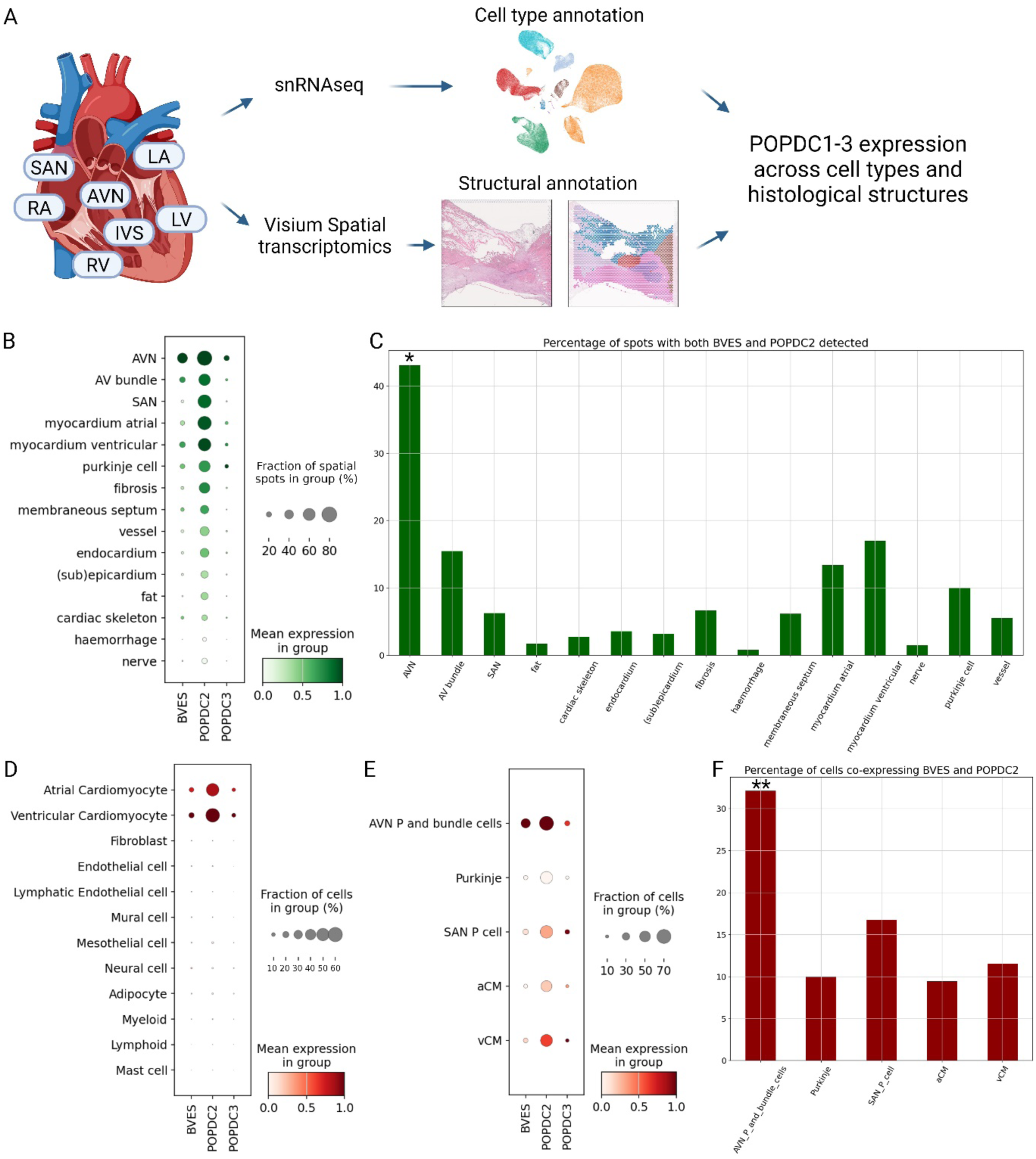
Expression of POPDC1-3 in human hearts. **(A)** Overview of single-nucleus and spatial transcriptomics data analysis from a previously published human heart cell atlas^14^ **(B)** Spatial transcriptomics (Visium) analysis of *POPDC1 (BVES), POPDC2* and *POPDC3* expression across different anatomical regions and histological microstructures in adult human hearts. The anatomical site sampled included AVN, SAN, left ventricle free wall, left ventricular apex, interventricular septum, left atrium and right atrium. **(C)** Percentage of spatial spots where co-expression of both POPDC1/BVES and 2 was detected are shown for each histological feature. * p<0.0001 from Chi square test comparing AVN vs all other cardiomyocyte-rich structures (AV bundle, SAN, myocardium atrial, myocardium ventricular and Purkinje cells). **(D)** *POPDC* family genes expression across cell types in adult human heart profiled by snRNAseq (10x Genomics). **(E)** *POPDC1/BVES, 2* and *3* gene expression in cardiomyocyte cell states in adult human hearts. **(F)** Percentage of cells which co-express *POPDC1/BVES* and *POPDC2* in the same single cardiomyocyte. ** p<0.0001 for Chi square test AVN P and bundle cells vs all other cardiomyocytes. Figure created with BioRender.

To validate these findings and to explore *POPDC1* and *2* co-expression within the same cell, we analyzed single-nucleus RNA sequencing (snRNAseq) data from 8 regions of adult human hearts including the conduction system. *POPDC2* expression was almost entirely restricted to atrial and ventricular cardiomyocytes compared to other cell types (**Figure 5D**).^14^ Other genes from the POPDC family i.e. POPDC1 and POPDC3 also showed a preferential expression in cardiomyocytes but their expression was less prevalent than POPDC2. The snRNAseq object was then sub-set to cardiomyocytes only and expression of POPDC1-3 interrogated. POPDC2 was expressed in all cardiomyocyte cell states while POPDC1 showed the highest expression in the pacemaker cells (P cells) of the AV node and the AV bundle cells (**Figure 5E**). The co-expression of POPDC1 and 2 within the same single-cell was most prevalent in AV node P cells and AV bundle cells which concurs with the spatial transcriptomic data and the observed predisposition to AV node disease in patients with POPDC2 loss-of-function variants (**Figure 5F**). We then also assessed the cardiac expression of *POPDC2* in mice utilizing previously published^15^ single-cell RNA sequencing data of the sinus node and AV node/His region, dissected from embryonic day 16.5 mouse hearts. Consistent with the phenotype observed in patients, *POPDC2* was expressed in cardiomyocytes as well as sinus node and AV node/His region cells (**Supplemental Figure 6**), corroborating observations from previous studies in mice^5,38^, and our observations reported here in human hearts (**Figure 5A-F**).

### Heterozygote carriers of rare *POPDC2* variants do not display characteristics of the recessive *POPDC2* syndrome at the population level

The recent availability of population-level cohorts with both clinical as well as whole-genome sequencing and well imputed array genotyping data now provide the opportunity for orthogonal validation of genetic findings through complementary population-level analysis. We investigated whether heterozygote carriers of the *POPDC2* variants found in families A-D (i.e. p.Q172-Y176delinsH, p.R263H, p.R263C, p.W188X and p.L37SfsTer20) showed (sub)-clinical manifestations of the recessive *POPDC2* syndrome. We searched their association with (1) AV block, (2) bradycardia, (3) cardiac arrest, (4) HCM, (5) myocarditis, (6) pacemaker implantation, (7) sinus node dysfunction and (8) heart rate. We detected 426 heterozygous carriers of the five familial *POPDC2* variants (p.Q172-Y176delinsH, n=62; p.R263H, N=34; p.R263C, n=156; p.W188X, n=162; and p.L37SfsTer20, n=12) among 1,089,031 participants in the included biobanks, but no homozygotes or compound heterozygotes. None of the variants showed statistical association with (sub)-clinical outcomes after Bonferroni correction, suggesting that heterozygous family members are unlikely to develop clinical manifestations and therefore might not necessitate clinical follow-up (**Supplemental Table 4**). We then conducted genetic burden analyses, wherein rare variants in *POPDC2* are aggregated, with the phenotypes above and muscular dystrophy as this is a clinical hallmark of recessive syndromes associated with the 2 other members of the POPDC family (i.e., *POPDC1*, and *POPDC3*). We found a significant association with bradycardia after Bonferroni-adjustment (P <1.4 ×10^-3^) for the following variant sets: LOFCADD (P=3.3×10^-4^, Odds ratio (OR) 1.6, LOF and missense when predicted deleterious with CADD phred score ≥ 25) and LOF1MISID (P=2×10^-4^, OR 1.59, LOF and missense when predicted deleterious by at least one of the following prediction methods: MetaSVM, MetaLR or CADD phred score ≥ 25, **Supplemental Table 5**). Although no association with a clinically relevant phenotype was detected an association with slow heart rate was seen in heterozygous carriers in the general population. Whether this has clinical relevance remains to be explored.

## DISCUSSION

We identified bi-allelic variants in *POPDC2* in 4 families that presented with a phenotypic spectrum consisting of sinus node dysfunction, AV conduction defects and variable HCM. Using homology modelling we show that the identified *POPDC2* variants are predicted to diminish the ability of POPDC2 to bind cAMP. In *in vitro* electrophysiological studies demonstrated that, while co-expression of WT POPDC2 with TREK-1 increased TREK-1 current density, POPDC2 harboring variants found in the patients failed to increase TREK-1 current density. scRNA-seq from human hearts demonstrated that co-expression of POPDC1 and 2 was most prevalent in AV node, AV node pacemaker and AV bundle cells. Sinus node cells expressed POPDC2 abundantly, but expression of POPDC1 was sparse. Together, these results concur with predisposition to AV node disease in humans with loss-of-function variants in *POPDC1* and *POPDC2* and presence of sinus node disease in *POPDC2* but not in *POPDC1* related disease in human. Our findings provide evidence for *POPDC2* as the cause of a novel Mendelian autosomal recessive cardiac syndrome.

Several observations support the causality of the identified *POPDC2* variants (p.Q172-Y176delinsH, p.R263H, p.R263C, p.W188X and p.L37SfsTer20). (1) The cardiac arrhythmia phenotype of the patients harboring these variants in the homozygous or compound heterozygous state is similar to observations of sinus pauses and bradycardia that were made in mice lacking *Popdc2*,^5^ and to AV block observed in zebrafish after morpholino knockdown of *popdc2*.^6^ (2) The variants affect highly conserved residues and are predicted damaging by multiple *in silico* prediction tools. (3) The variants are extremely rare in gnomAD^12^ and were not found in homozygous state. (4) A change affecting the paralogue residue of p.R263H (Family B) and p.R263C (Family C) in *POPDC3*, p.R261Q, has been associated with muscular dystrophy.^39^ The dimeric homology model of POPDC2 predicted all disease variants we identified in the homozygous or compound heterozygous state to critically affect cAMP binding. One variant leads to the substitution of five residues with a single histidine residue (p.Q172-Y176delinsH, **Figure 2B),** and another two (p.W188X/p.L37SfsTer20, **Figure 2C**) are expected to produce a truncated protein, all of which are expected to critically affect the ability of POPDC2 to bind cAMP.^26^ While the p.R263H and p.R263C variants are not predicted to disrupt protein folding, they are expected to eliminate a key interaction predicted to stabilize cAMP binding.^40^ Based on these predictions, we hypothesize that cAMP binding is critical for POPDC2 function, and when affected causes disease.

Although *POPDC2* has been proposed as a susceptibility gene for cardiac conduction disease in humans by Rinné et al.^41^, no Mendelian recessive disorder has been linked to this gene so far. In the study by Rinné et al.^41^, a heterozygous nonsense variant in *POPDC2* (p.W188X; rs144241265, allele frequency of 1.70×10^-4^ among European individuals in gnomAD^12^) was identified in the heterozygous state in a monozygotic twin pair presenting with AV block and in an unrelated family with a mother and son both diagnosed with 1^st^ degree AV block (i.e. prolongation of the PR-interval on the ECG). The twin pair inherited the variant from the unaffected mother, indicating that it does not fully explain the phenotype observed in the two siblings. In support of this, single-variant analysis that we conducted in 162 carriers of the p.W188X variant in more than 1 million individuals from the general population did not show an association with sinus node nor AV node conduction disease in heterozygous state. Thus, while the findings from Rinné et al.^41^ are interesting and support our findings, they do not establish *POPDC2* as a (recessive nor dominant) Mendelian cause of cardiac conduction disease in human. Based on our findings, we recommend the inclusion of *POPDC2* in clinical genetic testing panels for patients presenting with unexplained sinus node dysfunction, AV conduction defects with or without HCM. Using population-level genetic data of more than 1 million individuals we showed that none of the variants were associated with clinical outcomes in heterozygous state, suggesting that heterozygous family members are unlikely to develop clinical manifestations and therefore might not necessitate clinical follow-up.

Bi-allelic variants in *POPDC1* and *POPDC3* have been associated with muscular dystrophy, with and without CCD, respectively,^4,5,20^ while the patients reported here presented with isolated cardiac disease. Specifically, none of the patients reported muscular weakness, atrophy or cramps. Furthermore, muscle biopsy in the proband from Family A did not show evident signs of muscle disease. Also, in contrast to *POPDC1* and *PODPC3* patients, normal serum creatine kinase levels were found in the patients reported here. While we did not detect a muscular phenotype in the patients, currently aged 22-50 years, we cannot exclude subtle or age-dependent expression of muscular defects in *POPDC2*-related disease. Although all three POPDC proteins are expressed in both skeletal and cardiac muscle, differences in levels of expression between the POPDC proteins might, in part, determine phenotypic expression. Indeed, POPDC2 is predominantly expressed in cardiac tissue whereas POPDC3, which presents with isolated muscular dystrophy, has a predominant expression in skeletal muscle.^42^

Western blot analysis and immunostaining of POPDC1 and POPDC2 in muscle biopsies obtained in the proband from Family A showed significant reduction of the expression of both POPDC1 and POPDC2. These findings are in line with previous studies that suggested that stability and/or membrane trafficking of the POPDC1–POPDC2 complex is impaired by variants in each of the two proteins.^36,37^ Using spatial transcriptomics and scRNA-seq from human hearts we showed that co-expression of POPDC1 and 2 was most prevalent in AV node, AV node pacemaker and AV bundle cells. On the other hand, in the sinus node, POPDC2 was abundantly expressed, but expression of POPDC1 was sparse. Together, these results concur with predisposition to AV node disease in humans with loss-of-function variants in *POPDC1* and POPDC2 and presence of sinus node disease in *POPDC2* but not in *POPDC1* related disease in human.

POPDC proteins are established interacting partners of the potassium channel TREK-1, which is known to underlie a background potassium current and is highly expressed in the sinus node.^33^ Co-expression of TREK-1 with any of the three POPDC proteins leads to an increase in TREK-1 current, a process modulated by the level of cAMP.^3,5^ Furthermore, cardiac-specific TREK-1 deficient mice display a sinus node phenotype characterized by bradycardia with frequent episodes of sinus pauses, partially resembling the phenotype in the patients with *POPDC2* variants presented here.^33^ In an effort to shed light on the electrophysiological mechanism by which the variants in POPDC2 lead to bradycardia we therefore conducted co-expression studies of WT and mutant POPDC2 with TREK-1. In vitro, both *POPDC2* variants tested failed to increase TREK-1 current and, by virtue of observations of bradycardia in TREK-1 deficient mice, these variants would be expected to be associated with bradycardia.

Notwithstanding the clear observation of bradycardia in TREK-1 deficient mice, how loss of background potassium current, causing an increase in diastolic net inward current, leads to bradycardia and sinus pauses is unclear. Our *in silico* modeling studies showed that a 41% reduction of TREK-1 current, simulating the effect of the variants in *POPDC2,* leads to an increase in diastolic depolarization rate and spontaneous firing. These findings are in agreement with experiments using isolated sinus node cells of TREK-1 deficient mice and computer simulations using a rabbit sinus node cell model.^33^ In a murine cardiac muscle cell line (HL-1), a stop variant in *popdc2* associated with TREK-1 reduction, the maximum diastolic potential (MDP) was depolarized and the action potential upstroke velocity was reduced.^41^ In addition, a slower spontaneous firing rate was observed.^41^ In contrast, another study that examined loss of TREK-1 channel function in HL-1 cells, showed an increase in spontaneous firing rate.^43^ Bradycardia may also be induced via changes in excitability of atrial cardiomyocytes surrounding the sinus node. In our simulated human atrial cell, reduction in TREK-1 current density slightly depolarized the MDP and increased the APD, consistent with findings in rat ventricular myocytes,^44^ but the excitability was hardly affected.

Thus, while collectively these data support a role for TREK-1 in cardiac pacing and a causal effect of *POPDC2* variants through modulation of TREK-1 current, the exact cellular electrophysiological mechanism remains unclear. Differences in the cellular models used cannot be excluded. Furthermore, the differences in effects of TREK-1 deficiency observed *in vivo* and *in vitro* in TREK-1 deficient mice suggest that other factors (such as altered sympathetic or parasympathetic stimulation *in vivo*) also contribute to the bradycardia observed at baseline in these mice.^33^ While an effect through modulation of sodium channel function could be postulated, no such effect was observed in our patch-clamp studies.

Although cardiac arrhythmias in patients with recessive *POPDC2* variants are consistent with observations in mice^5^ and zebrafish^6^, the role of *POPDC2* in cardiac hypertrophy remains unexplained. One of the potential mechanisms underlying cardiac hypertrophy in *POPDC2* patients is modulation of TREK-1. Since TREK-1 is activated by biomechanical stretch, the role of TREK-1 in cardiac responses to chronic pressure was recently studied using transverse aortic constriction.^45^ Notably, while no clear structural cardiac differences were seen at baseline, mice lacking TREK-1 exhibited an exaggerated pressure overload-induced concentric hypertrophy with preserved systolic and diastolic cardiac function, compared to wild-type mice.^45^

Patient 6 from Family D was diagnosed with bradycardia resulting in an arrest and 1^st^ degree AV block during an episode of fulminant myocarditis. We therefore cannot exclude a causal role of myocarditis in this case. However, (1) the conduction phenotype fits with the phenotypic characteristic of the other five patients we report here, (2) the patient carried bi-allelic truncating variants in *POPDC2* likely to result in complete loss of function, which is a known mechanism for disease in animal models, (3) the variants found in compound heterozygosity in this case were never seen together in 141,456 individuals from gnomAD, (4) gnomAD contains 70 predicted loss-of-function *POPDC2* variants; none of them occurs in the homozygous state, (5) knock-in mice of the p.W188X variant (found in compound heterozygosity in Family D) displayed stress-induced sinus bradycardia and pauses. Presentation with (recurrent) myocarditis-like episodes has been clearly established for arrhythmogenic cardiomyopathy (ACM).^46^ Among 560 probands and family members with ACM, Bariani et al. reported an episode resembling myocarditis (i.e.’hot phase’) in 23 cases (5%), particularly in pediatric patients and carriers of desmoplakin gene variants.^47^

Furthermore, in a population-based cohort of 336 consecutive patients with acute myocarditis, a significant enrichment of pathogenic variants in genes associated with dilated or arrhythmogenic cardiomyopathy was found (8%) in comparison with controls (<1%, *P*=0.0097). The question remains whether the myocarditis exposed the underlying *POPDC2* related conduction disease or whether myocarditis is part of the *POPDC2* phenotypic spectrum. While we find it unlikely, compound heterozygosity of the truncating *POPDC2* variants, might have been irrelevant in this case and the phenotype could be fully the consequence of myocarditis.

In summary, our findings associate *POPDC2* with a novel Mendelian autosomal recessive cardiac syndrome consisting of sinus node dysfunction, AV conduction defects and variable HCM.

## Data Availability

The pdb coordinates for the POPDC2 homology models are available on request from the authors. All data produced in the present study are available upon reasonable request to the authors

## ACKNOWLEDGEMENTS

We thank the families for their participation and collaboration. N.L. is supported by the Dutch Research Council (ZonMW VENI and Off-road). C.R.B., A.V.P. and N.L. acknowledge the support from the Dutch Heart Foundation (CVON 2018-30 PREDICT2 and CVON2014-18 CONCOR-GENES to C.R.B.) and the Netherlands Organization for Scientific Research (VICI fellowship, 016.150.610, to C.R.B.). This work was supported in part by the NIH awards R35GM128666 (M.V.A.) and T32GM092714 (F.Z.B.), a Sloan Research Fellowship (M.V.A.), and an American Heart Association Fellowship 23PRE1019634 (L.W.). This study makes use of data generated by the DECIPHER community. A full list of centres who contributed to the generation of the data is available from http://decipher.sanger.ac.uk and via email from. Funding for the project was provided by the Wellcome Trust. Firth, H.V. et al (2009). DECIPHER: Database of Chromosomal Imbalance and Phenotype in Humans using Ensembl Resources. Website: https://www.deciphergenomics.org/. This project has been made possible in part by the Chan Zuckerberg Foundation (2019-202666) to M.N. and the British Heart Foundation and Deutsches Zentrum fur Herz-Kreislauf-Forschung (BHF/DZHK: SP/19/1/34461) to M.N. M.N. and L.M. were supported by the Rosetrees Trust Intermediate Project Grant (PGS23/100028) and NIHR Imperial BRC Cardiovascular Pilot Project Grant. L.M. was further supported by British Heart Foundation Clinical Research Training Fellowship (FS/CRTF/23/24444), British Society for Heart Failure Research Fellowship and Alexander Jansons Myocarditis UK. We thank Drs. Delphine Bichet and Florian Lesage (Universite de Nice Sophia Antipolis, France) for sharing the hTREK-1a plasmid. We thank Drs. Mohamed Hosny and Magdi Yacoub (Aswan Heart Centre, Magdi Yacoub Foundation, Egypt) for reviewing the phenotype of patients recruited at their center. We thank Sara Teichmann for sharing data on scRNA seq in human hearts.

## ETHICS DECLARATION

The study protocol was approved by the Amsterdam University Medical Center Research Ethics Committee and the local Institutional Review Boards of contributing centers. Signed informed consent was obtained from the patients or their parents.

## AUTHOR CONTRIBUTIONS

**Conceptualization:** N.L., C.R.B.; **Data curation:** N.L., M.N., A.M.C.V. P.G., R.T., A.V.P., E.M.L. P.A.Z., M.A., M.A., Y.A., L.M., K.K., J.G., S.R., H.B., H.U., C.E., B.A., M.T.B., M.C., H.K.J., D.A.C, C.T., K.F., P.M.T., K.B., T.M.K., L.S., B.G.W., J.M., F.F., G.P., D.R., J.P.T., M.N., M.A.V., I.C., A.A.M.W., R.W., S.A.C.; **Investigation:** N.L., M.N., F.Z.B., L.W., V.T., L.M., K.D., R.Z., L.M., J.L.S., D.U., A.M.A.M, K.K., J.C., I.E.Z., E.M.A.A., M.R., G.S., E.V.I., H.H., D.F.G., A.T.S., K.S., L.N., K.U.K., S.R.O., E.S., O.B.V.R., F.F., G.P., D.R., J.P.T., R.W., A.O.V; **Visualization:** N.L., M.N., F.Z.B., L.W., L.M., K.D., K.K., M.R., S.Z., R.W.; **Writing-original draft:** N.L., M.N., C.R.B., R.W., A.O.V.; **Project administration:** N.L., C.R.B., P.G.P., L.B.; **Writing-review & editing:** N.L., M.N., R.W., A.O.V., C.R.B.; **Supervision:** N.L. All authors read, revised, and approved the final manuscript.

