## Supplemental Text and Figures for "Biallelic variants in *POPDC2* cause a novel autosomal recessive syndrome presenting with cardiac conduction defects and variable hypertrophic cardiomyopathy"

Nicastro et al.

**Table of contents**

**Supplemental Methods**
**Supplemental Notes**
**Supplemental Figure 1.** Holter registration of the proband from Family A.
**Supplemental Figure 2.** Structure information for POPDC2 homology modeling.
**Supplemental Figure 3.** Effects of WT (wild-type) and mutant POPDC2 on sodium current ($I_{Na}$) expressed in HEK-293 cells.
**Supplemental Figure 4.** Expression level of mutant POPDC2 is not reduced compared to wild type.
**Supplemental Figure 5.** Western blot analysis of POPDC1 and POPDC2 in protein lysates from muscle biopsies.
**Supplemental Figure 6.** Expression of POPDC proteins in the developing mouse heart.

### Supplemental Methods

#### Whole exome sequencing and candidate screening of *POPDC2*

**Family A:** Genomic DNA was isolated from peripheral blood. Whole exome sequencing was conducted on the proband, both parents and two unaffected siblings from Family A. Exome capture was performed using the SeqCap EZ Human Exome Library v3.0 (Roche NimbleGen) and sequencing was performed on an Illumina HiSeq2500. The read alignment to GRCh37 (hg19) and the variant calling were done with a pipeline based on the Burrows-Wheeler Aligner[1] and the Genome Analysis Toolkit.[2] Variant annotation and prioritization were done with Cartagenia Bench Lab next-generation-sequencing (Agilent Technologies). Variants located in exon sequences and in flanking intron boundaries (≈6 nucleotides at exon-intron junctions) that had a minor allele frequency of <1% in control and in-house databases were included. In line with the expected transmission in the pedigree, variants that fitted with a de novo or recessive mode of inheritance were further analyzed. Validation of the *POPDC2* variant found in Family A was conducted by Sanger sequencing.

**Family B:** In cohort 1, 2 and 4, Sanger sequencing of the entire coding region of *POPDC2* and exon-intron boundaries was conducted using PCR primers listed in **Supplemental Table 1.** In cohort 3, next-generation sequencing of *POPDC2* was conducted as follows: *POPDC2* gene enrichment was performed through a whole gene direct PCR amplification using the following primers: POPDC2-WG-FW-P: 5'-GTGCTACTTTCCCTTGGGACTA-3' and POPDC2-WG-RV-P: 5'-TTGTAAGTTGAATGTCACAGCC-3'. The following PCR mix was used: 1 µL of oligo mix, 25 µL of mastermix (LongAmp® Hot Start Taq 2X Master Mix - New England Biolabs, USA), 5µL of betaine, 14 L of molecular grade water and 5 µL of DNA. *POPDC2* amplicons span a region of 18914bp (chr3:119360745-119379658), allowing read of the full coding region plus the 5'UTR promoter region. Patient's *POPDC2* amplicons were subjected to a next-generation sequencing protocol which included: DNA shearing in Covaris E220 (Covaris, France) and library preparation (NEXTFLEX Rapid DNA-Seq Kit – Perkin Elmer Inc, USA). Barcoded *POPDC2* enriched libraries underwent multiplexed NGS sequencing in a HiSeq 1500 instrument (Illumina Inc, USA). Generated raw data was processed through a bioinformatic pipeline covering: bcl files demultiplexing, duplicate reads removal, mapping (hg19), indel realignment, base-quality recalibration, variant calling and variant annotation. Validation of the *POPDC2* variant found in Family B was conducted by Sanger sequencing using the polymerase chain reaction (PCR) primers listed in **Supplemental Table 1**.

**Family C**: DNA was isolated from peripheral blood. Exome sequencing was performed using NimbleGen SeqCap EZ Medexome and sequencing was performed on an Illumina NovaSeq 6000. The read alignment to GRCh37 (hg19) and the variant calling were done with a pipeline based on the Burrows-Wheeler Aligner and the Genome Analysis Toolkit. Variant annotation and filtering were performed in Varseq v. 2.2.2. Variants located in exon sequences and in flanking intron boundaries (±20 nucleotides at exon-intron junctions) that had a minor allele frequency of <1% in GnomAD. In the family, there were no similar cases of cardiac disease in early age. Their parent was known with atrial fibrillation at an older age but showed no signs of bradycardia. In line with the expected transmission in the pedigree, variants that fitted with a recessive mode of inheritance were further analyzed.

**Family D:** Using genomic DNA from the proband and parent, the exonic regions and flanking splice junctions of the genome were captured using the IDT xGen Exome Research Panel v2.0 (Integrated DNA Technologies, Coralville, IA). Massively parallel (NextGen) sequencing was done on an Illumina system with 100bp or greater paired-end reads. Reads were aligned to human genome build GRCh37/UCSC hg19 and analyzed for sequence variants using a custom-developed analysis tool. Reported variants were confirmed, if necessary, by an appropriate orthogonal method in the proband and selected relatives. Additional sequencing technology and variant interpretation protocol has been previously described.[3] The general assertion criteria for variant classification are publicly available on the GeneDx ClinVar submission page (http://www.ncbi.nlm.nih.gov/clinvar/submitters/26957/).

**Study cohorts**

**deCODE**: All Icelandic genotyped participants that donated samples signed a written informed consent allowing the use of their samples and data in research projects at deCODE genetics. Data were anonymized and encrypted by a third-party system.[4] The study was approved by the Icelandic Data Protection Authority and the National Bioethics Committee of Iceland (VSN-15-057 and VSN-17-142-V5). Icelandic phenotype data were sourced from, Landspitali - The National University Hospital of Iceland, Primary Health Care Clinics of the Capital area and Akureyri Hospital, the regional hospital in North Iceland.

**UK Biobank:** The UK Biobank data were obtained under application number 56270. All participants provided an informed consent for the use of genotype data and link to electronic health records (EHR). Phenotypes were provided by the UK Biobank from its assessment centre, hospital records and general practitioner clinics. Participants enrolled in the study between 2006 and 2010 throughout the UK and were aged 38-65 years at recruitment. Only Caucasian British and Irish participants were used in this study.

**Copenhagen Hospital Biobank (CHB) and The Danish Blood Donor Study (DBDS)**: The Danish data were a combination of the Copenhagen Hospital Biobank study on personalized medicine in oral-cardiometabolic health (Copenhagen Hospital Biobank Oral-Cardiometabolic Health Cohort, CHB-OCHC) and the Danish Blood Donor Study (DBDS) I+II. Participants in the CHB-OCHC have all been informed about the use of their samples for research and the study was approved by the Zealand Region Committee on Health Research Ethics and the Capital Region Data Protection Office (SJ-989 and P-2022-913). DBDS participants have all consented to research on genetics and age-related diseases and the genetic studies were approved by the National and Zealand Regional Committees on Health Research Ethics as well as the Capital Region Data Protection Office (NVK-1700407, SJ-470, and P-2019-99). Phenotype data comes from Danish national health care registers.

**USA datasets:** Subjects from Utah, USA, derived from Intermountain Healthcare, a healthcare system of 24 hospitals and 160 clinics, using diagnoses between 1993-2022. The dataset was sourced from HerediGene, a general population study, and the INSPIRE Registry of individuals with heart disease. This study has been approved by the Intermountain Healthcare Institutional Review board and all participants have provided written informed consent.

### Genotyping

**deCODE**: The genome of the Icelandic population was characterized by whole-genome sequencing (WGS) of 63,460 Icelanders using Illumina standard TruSeq methodology to a mean depth of 35x (SD 8x) with subsequent long-range phasing,[5] and imputing the information into 173,025 individuals chip-genotyped employing multiple Illumina platforms.[6]

**UK Biobank:** The UK Biobank dataset consists of 428,503 whole-genome sequenced individuals of British or Irish ancestry, determined by principal component analysis. Sequencing was performed at deCODE genetics and Wellcome Trust Sanger Institute [7,8]

**CHB/DBDS/Intermountain:** We created a reference panel of 51,271 whole-genome sequenced individuals of multiple ancestries to a median coverage of 20x at deCODE genetics (of which 10,828 were Danes and 23,288 were Americans of European ancestry from Utah). We used this panel to impute the genotypes of up to 487,356 chip-typed individuals in the Danish dataset (DK), and 138,006 chip-typed individuals from the Intermountain dataset (US).Variant calling was performed using GraphTyper v2.6[9] and chip data was phased using SHAPEIT4.[10]

### Association analyses

Association testing of single variants and the burden of loss-of-function variants in *POPDC2* was conducted under an additive model using logistic regression for case/control phenotypes and BOLT-LMM for quantitative phenotypes.[11] Phenotype status was the dependent variable and carrier status (coded 1 for a minor allele for each specific variant in the single variant analysis, and for at least one loss-of-function variant in the gene burden analysis) was the independent variable. For individuals without whole-genome sequencing data in the burden analysis, imputed variants with imputation information > 0.7 were used. Covariates in each cohort were:

- Iceland: Sex, county of birth, current age/age at death, chip type availability, whole-genome sequencing data availability, and an indicator function for the overlap of lifetime with the time span of phenotype collection.
- Denmark: Sex, current age/age at death, whole-genome sequencing data availability, 12 principal components
- USA: Sex, current age/age at death, whole-genome sequencing data availability and 4 principal components
- UK: Sex, current age/age at death, whole-genome sequencing data availability for the individual, and 20 principal components

Standard errors were calculated in the following way:

For a *P*-value smaller than 1 we calculate the standard error as follows:

$$P \ = \ 2\Phi(z) = 2\Phi\left(\frac{\beta}{\alpha}\right).$$

Solving for *σ* gives

$$\sigma = \frac{\beta}{\Phi^{-1}\left(\frac{P}{2}\right)}$$

If *P* = 1, then and the above method breaks down. In this case we use data from other markers to estimate the relationship between allele frequency (*f*) and imputation information (*I*) and σ as follows:

$$Var(\beta) = \sigma^2 \propto \ \frac{1}{N} f(1-f) \propto \ \frac{1}{I} f(1-f)$$

Sample size (*N*) is proportional to imputation information (I), basing the analysis on the same set of individuals. Therefore, if we fit the following linear model:

$$\log(\sigma^2) = \gamma_1 + \gamma_1 \log(I) \ \ + \ \gamma_f \log\big(f(1-f)\big)$$

for a subset of 100,000 markers spread over the genome with MAF ranging close to uniformly between 0.1 and 50% and info between 0.9 and 1 and pick the subset of markers with *P* < 0.9 then we can predict σ for a marker with *P* close to 1.

**Supplemental Note 1.** Clinical description of patient 1-6 from families A-D

**Family A, patient 1**

The proband from Family A presented at the age range of 11-15 years. His parents are first cousins (**Figure 1A, individual II-3 in the pedigree**). He presented to the emergency department with palpitations associated with mild 'flu-like symptoms' and reported experiencing multiple intermittent episodes of chest pain and palpitations during both rest and exercise in the preceding 8 years. He never had syncope and was otherwise healthy. The patient did not display dysmorphic features or extracardiac abnormalities. He had normal serum creatine kinase (CK) levels and no muscular phenotype was observed during neurological evaluation. The electrocardiograms (ECGs, **Figure 1B**) at presentation showed sinus rhythm with sinus pauses and junctional and ventricular escape beats, as well as type 1 (Wenckebach) second-degree AV block. There were frequent unifocal premature ventricular contractions (PVCs), sometimes in bigeminy. In-hospital and ambulatory (Holter) cardiac monitoring showed marked sinus bradycardia (33 beats per minute, bpm) with multiple sinus pauses up to 3 seconds mostly during sleep (**Supplemental Figure 1**), as well as a high burden (13% of total beats) of ventricular arrhythmia, including rapid monomorphic non-sustained ventricular tachycardia up to 234 bpm, and paroxysmal atrial flutter/fibrillation with slow ventricular rate. Exercise test (X-ECG) showed normal acceleration of sinus rate with normal AV conduction up to 181 bpm and episodes of type 1 (Wenckebach) second-degree AV block in the recovery phase. Trans-thoracic echocardiography revealed marked septal cardiac hypertrophy (23 mm, Z-score30: 16.43, height, 160 cm; weight, 49 kg) and normal left ventricular systolic function. Systolic anterior motion of the mitral valve was not observed, and there was no left ventricular outflow tract obstruction. Cardiac magnetic resonance imaging (MRI) confirmed the marked asymmetric left ventricular hypertrophy (maximal septal thickness 23 mm, **Figure 1B**) with normal systolic function, and demonstrated the presence of myocardial fibrosis (late gadolinium enhancement) in the basal infero-septal region where the hypertrophy was the most prominent. Because of the ventricular arrhythmias in the context of underlying severe HCM at young age, a transvenous implantable dual-chamber cardiac defibrillator was implanted for primary prevention. The patient is currently 21-25 years old and no major cardiac events have occurred during follow-up. Clinical evaluation of his parents at the age 41-45 and 41-45 years, respectively, and his three siblings (age 0-5, 6-10 and 16-20 years) was performed (including ECG recording and echocardiography), which did not reveal any abnormality. Reevaluation of his youngest siblings at the age of 6-10 and 11-15 years (including ECG recording, X-ECG, Holter monitoring and echocardiography) did not show any signs of cardiac arrhythmia or cardiomyopathy.

**Family B, patient 2**

The proband from Family B presented at the age range of 21-25 years. His parents are first cousins (**Figure 1A, individual II-4 in the pedigree**). He initially presented with complaints of palpitations. He reported episodes during which he felt his heart suddenly beating considerably faster or slower than usual. He was otherwise healthy, and the physical examination was normal. He had no symptoms of myopathy. An ECG, Holter recording, and echocardiography were performed. The PQ time was at the upper range of normal (200 ms, normal range 120-200 ms). The Holter recording showed recurrent episodes of concurrent sinus bradycardia and type I (Wenckebach) second-degree AV block during sleep and while awake (**Figure 1B**). Echocardiography showed diastolic dysfunction and discrete interventricular septal hypertrophy (11 mm). An X-ECG and cardiac MRI were then performed. The X-ECG showed no abnormalities. The MRI showed asymmetric hypertrophy with a maximal wall thickness of 16 mm at the basal septum without signs of left ventricular outflow tract obstruction (left ventricular posterior wall thickness 8 mm). No late gadolinium enhancement was noted on the MRI. No other structural abnormalities were present and biventricular systolic ventricular function was normal. No history of cardiac conditions or complaints were known for the parents. A sibling of the patient had a rhythm disorder requiring pacemaker implantation before the age of 25 years (II-3). Relatives were unavailable for further genetic and clinical evaluation.

**Family C, patient 3-5**

The proband was a 46-50 year-old male referred for genetic reevaluation (**Figure 1A, Family C, individual II-3 in the pedigree**). At the age of 16-20 years, the patient had a cardiac arrest with asystole. The ECGs showed ST elevation in V2 and V3 and negative T waves in V3-V6 and in I, II and III. Coronary angiography and echocardiogram were both normal. Electrophysiological examination showed clear signs of bradycardia, first and second-degree AV block (Wenckebach), non-inducible for VT, as well as episodes of sinus arrest. Planar myocardial perfusion imaging and myocardial biopsy both indicated possible myocarditis. A dual chamber (atrioventricular) pacemaker was implanted on the indication bradycardia induced cardiac arrest. Repeated echocardiography at the age of 31-35 years showed onset of dilatation of the right ventricle with normal tricuspid annular plane systolic excursion (TAPSE, indicative of normal right ventricular function). Reassessment of the myocardial biopsy indicated fatty tissue and fibrosis suggestive of arrhythmogenic cardiomyopathy, but not reaching current guidelines for ARVC diagnosis. Combined with an outpatient cardiac monitoring showing non-sustained ventricular tachycardia, arrhythmogenic cardiomyopathy was suspected, and the pacemaker was upgraded to a dual-chamber implantable cardiac defibrillator (ICD). At a base rate of 50 bpm the pacemaker showed 82% atrial pacing and approximately 50% ventricular pacing. The pacemaker also showed some atrial high-rate episodes, SA-block and one incident (at the age of 46-50 years) with a short non-sustained VT. There was one reported monomorphic NSVT (4 beats, 225 bpm) documented on outpatient cardiac monitoring (**Figure 1B**). The ICD has never given any appropriate shock and there has been no sustained ventricular tachycardia. Genetic testing was previously performed including ~250 genes related to inherited cardiovascular disease. No pathogenic variants were detected. The proband had two siblings who were clinically evaluated due to proband's incidence of cardiac arrest. Both siblings showed signs of bradycardia on the electrocardiograms (ECGs).

At the age of 21-25 years his older siblings underwent clinical evaluation (**Figure 1A, Family C, individual II-2 in the pedigree**). A cardiac event recorder showed signs of sick-sinus syndrome (bradycardia, up to 5 seconds sinoatrial (SA) block), with no presyncope or syncope, and was treated with a pacemaker. The ECGs before implantation of pacemaker showed incomplete right bundle branch block. The echocardiography was described as normal. At the age of 41-45 years, a new echocardiogram showed dilated right and left atria. The pacemaker was programmed in AAI due to a dysfunctional ventricular electrode. At the last pacemaker readout, at a base rate of 30 bpm, the pacemaker showed 4.8% atrial pacing, without atrial or ventricular arrhythmia.

At the age of 21-25 the oldest sibling was clinically investigated (**Figure 1A, Family C, individual II-1 in the pedigree**). His ECG showed 1$^{st}$ degree AV block and 2nd degree AV-block (Type 1 and 2:1) with asymptomatic pauses up to 3.7 seconds (**Figure 1B**). The echocardiography was normal. A dual chamber pacemaker was implanted. Eight years later, a myocardial biopsy was performed which was described as normal. At the age of 41-45 years, he developed symptomatic atrial flutter, and ablation (right-side isthmus block) was performed. Only one episode of monomorphic non-sustained VT has been observed. The patient had echocardiography at a regular basis with at LVEF 60% and no dilatation. At the last pacemaker readout, at a base rate of 40 bpm, the pacemaker showed 0% atrial pacing and approximately 23% ventricular pacing, without atrial or ventricular arrhythmia.

None of the patients from Family C showed signs of hypertrophic cardiomyopathy. In addition, to our best knowledge, none of the siblings had any chronic neuromuscular complains or known neuromuscular disease, besides that the individual II-1 did experience severe muscular pain after heavy exercise as well as an episode of severe general muscular pain at the age of 41-45, which was interpreted as due to a viral infection. His CK levels were within normal ranges (measured 41-45 years old; 115 and 202 U/L (reference 50-400). Exome sequencing was conducted in all three siblings. A rare homozygous variant in POPDC2 was identified in all three siblings: NM_001369919.1:c.787C>T; NP_001356848.1:p.Arg263Cys.

In the family, there were no similar cases of cardiac disease at an early age. Their parent was known with atrial fibrillation at an older age but showed no signs of bradycardia. The parent is in general asymptomatic and experiences slightly sense of palpitations, lasting a few seconds, 2-3 times a week. The ECG shows incomplete RBBB. The echocardiogram showed LVEF of 60, with slightly restrictive contractility and borderline LV hypertrophy, without valvular disease. Long term ECG monitoring revealed two short incidents with non-sustained ventricular tachycardia (VT) and short episodes of supraventricular tachycardia at a rate of 170 bpm. Cardiac MRI showed LVEF 55% without late gadolinium enhancement. Cardiac-CT displayed an insignificant small plaque in the left anterior descending artery. In general, the patient is very sensitive to bradycardia in her treatment with beta-blockers. Both parents were heterozygous for the variant in POPDC2. There was no known consanguinity in the family, but when asked there might have been a remote relative with the same family name. Her sibling was offered testing for the variant, but due to other diseases and old age she declined. One relative had atrial fibrillation at an early age (41-45 years old) and 3-degree AV block at the age of 86-90. He did not carry the variant. Another relative had possible episodes of atrial fibrillation in older age. He was heterozygous for the POPDC2 variant. None of his adult children were known with cardiac diseases.

**Family D, patient 6**

The proband is a 11-15-year-old male (**Figure 1A, Family D, individual II-1 in the pedigree)**. He started having episodes of chest pain for 2-3 days after which he presented to the emergency department (ED). He described the pain as sharp, stabbing with some shortness of breath, cough, and vomiting. His troponin was significantly elevated at 225,000 pg/mL (normal less than 21 pg/mL), there was diffuse ST segment elevation on the ECG (particularly in the inferior and lateral leads, I, II, AVF), and elevated aspartate aminotransferase (AST) of 549 U/L. A CT of the chest was performed to evaluate for pulmonary embolism and was read as negative. The echocardiography at presentation showed severe biventricular dysfunction. He was admitted to the cardiac intensive care unit and had initiation of inotropes. He was first started on non-invasive ventilation after which he was intubated. The chest X-ray was consistent with pulmonary oedema. Overnight, he had an episode of acutely hypotensive cardiac arrest that was not precipitated by an arrhythmia but was associated with bradycardia (HRs in 40s-60s). Associated with his first arrest he transitioned to ventricular tachycardia with pulse and was cardioverted and amiodarone was initiated. He was taken to the cardiac catheterization lab where he had a second arrest associated with profound hypotension, bradycardia, and collapse with pulseless electrical activity, requiring cardiopulmonary resuscitation and epinephrine. Hemodynamics showed markedly reduced cardiac index, markedly elevated left ventricular end-diastolic pressure and left atrial pressure. Angiography was negative for pulmonary embolism and coronary artery angiography negative for coronary event. He was treated empirically for myocarditis with intravenous immunoglobulin (IVIG) and steroids. Cardiac biopsy showed non-specific focal mild interstitial fibrosis of the endomyocardium Importantly, cardiac biopsies were negative for inflammation without histological evidence for myocarditis. Temporary pacemaker leads were placed for bradycardia. He continued to recover and underwent cardiac MRI. There was concern for extensive midmyocardial delayed contrast enhancement. A second cardiac MRI was obtained which confirmed a significant amount of fibrosis and inflammation circumferentially around the left ventricle. He was discharged home with a Zoll life vest (with the concern for fibrosis and possibility of ventricular arrhythmias). No arrhythmias have been recorded on the vest. Follow-up echocardiography showed moderate LV systolic dysfunction without dilatation or hypertrophy, and normal RV size and function.

**Supplemental Note 2.** p.R263C is not a founder variant in the Danish population

We hypothesized that p.R263C is a founder variant in the Danish population. However, the MAF of p.R263C in the Danish Copenhagen Hospital Biobank (n=487,356, MAF $1.48 \times 10^{-4}$) was comparable with the MAF in gnomAD V4.0 (n=807,162, MAF $1.13 \times 10^{-4}$). We then searched for p.R263C in a recent whole-genome sequencing dataset[12] consisting of young patients (<50years, n=226) receiving a pacemaker because of AV block, identified through Danish Pacemaker and Implantable Cardioverter Defibrillator Registry, to which all pacemaker implantation procedures in Denmark are reported (January 1, 1996 to December 31, 2015). None of the patients was homozygous for p.R263C nor were there any patient with bi-allelic variants in *POPDC2*. Together, these finding argue against p.R263C being a founder variant. Indeed, while there was no known consanguinity in the family, the parents noted that there might have been a remote relative with the same family name as the most likely explanation for homozygosity of p.R263C.

**Supplemental Figures**

**Supplemental Figure 1.** Holter registration of the proband from Family A

Ambulatory (Holter) cardiac monitoring showed marked sinus bradycardia (33 beats per minute, bpm) with multiple sinus pauses up to 3 seconds mostly during sleep (example tracing).

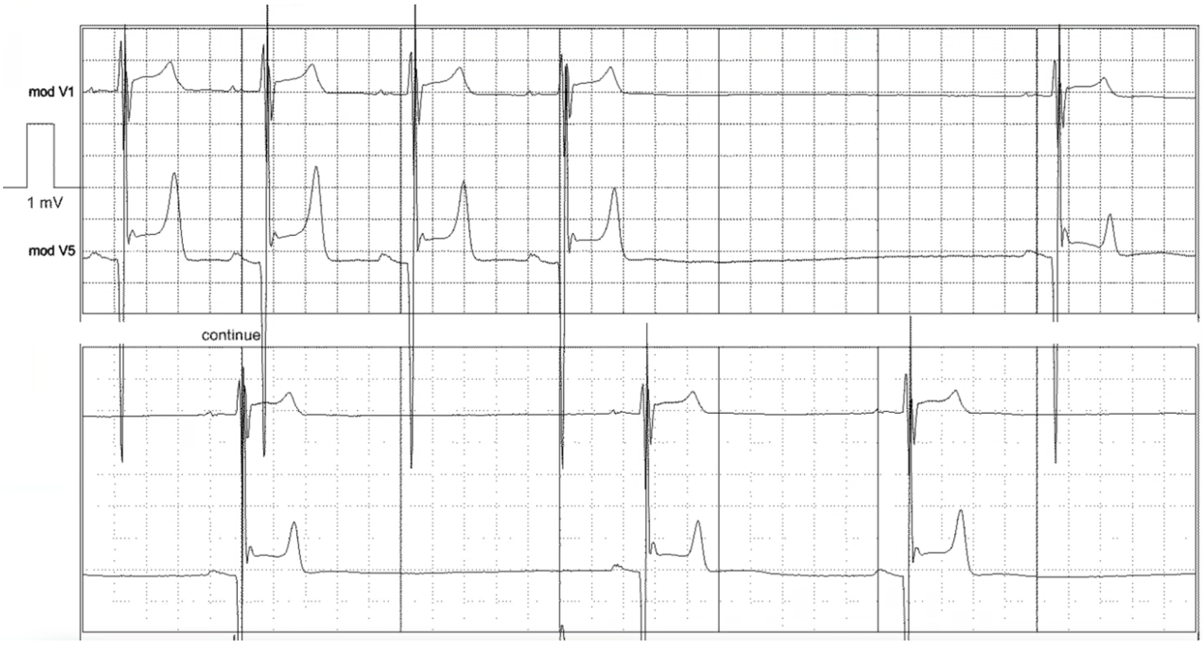

**Supplemental Figure 2.** Structure information for POPDC2 homology modeling. (**A**) Homology model of POPDC2 by AlphaFold Multimer, color coded by relative confidence level. **(B)** Homology model of POPDC2 Popeye domains by SWISS-MODEL with cAMP bound.

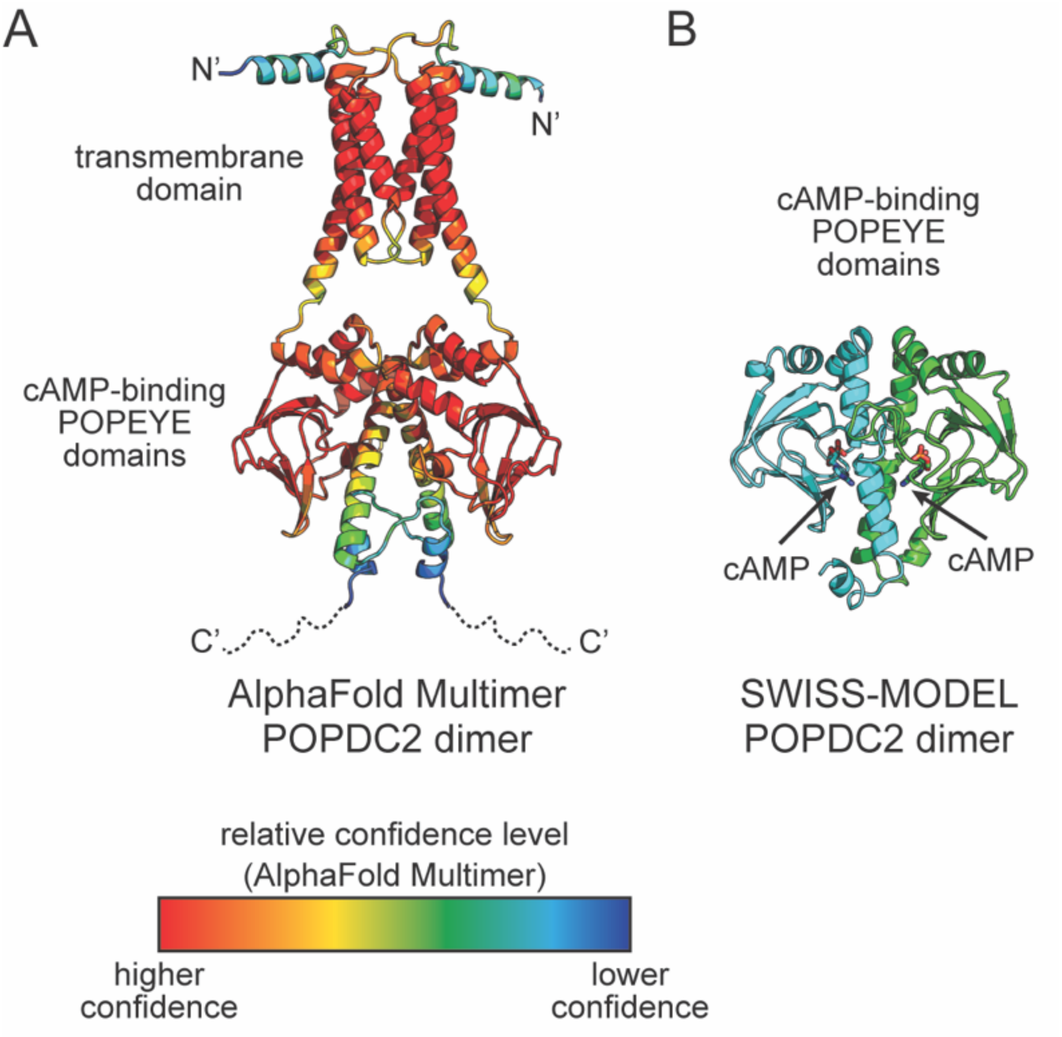

**Supplemental Figure 3.** Effects of WT (wild-type) and mutant POPDC2 on sodium current ($I_{Na}$) expressed in HEK-293 cells. (**A**) Current-voltage relationships of $I_{Na}$ in absence or presence of WT and mutant POPDC2. (**B**). $I_{Na}$ peak amplitude in absence or presence of WT and mutant POPDC2. (**C**) $V_{1/2}$ (left) and k (right) of $I_{Na}$ activation in absence or presence of WT and mutant POPDC2. (**D**) $V_{1/2}$ (left) and k (right) of $I_{Na}$ inactivation in absence or presence of WT and mutant POPDC2. One-way ANOVA testing did not reveal statistically significant differences in the $I_{Na}$ properties. **Abbreviations:** WT, wild-type; $V_{1/2}$, membrane potential for half-maximal (in)activation; k, slope factor.

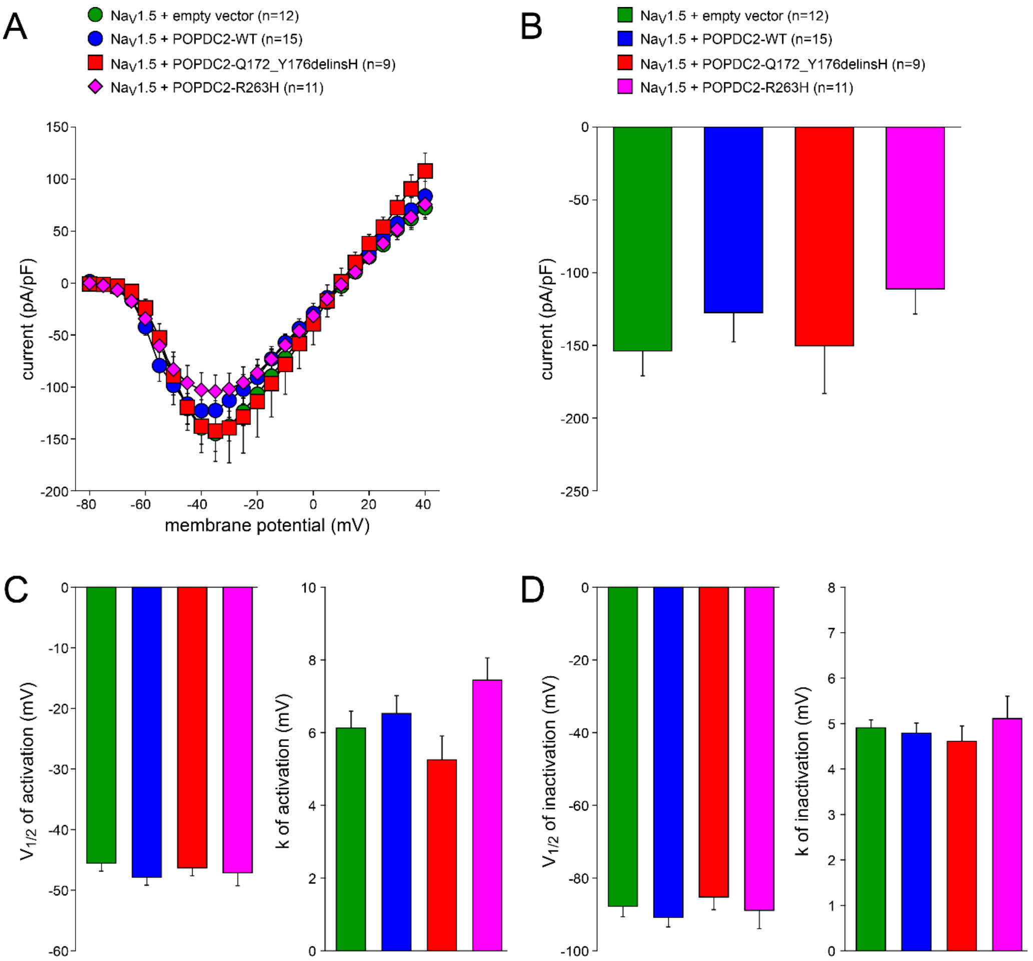

**Supplemental Figure 4.** Expression levels of mutant POPDC2 are not reduced compared to wildtype (WT). Each column contains samples from an independent transfection (n=3). Mutant POPDC2 protein expression (p.R263H and p.Q172_Y176delinsH) was comparable to wild type after normalization to GFP. Every transfection was monitored by qRT-PCR which showed a positive outcome of the transfection protocol. GAPDH is included as a loading control.

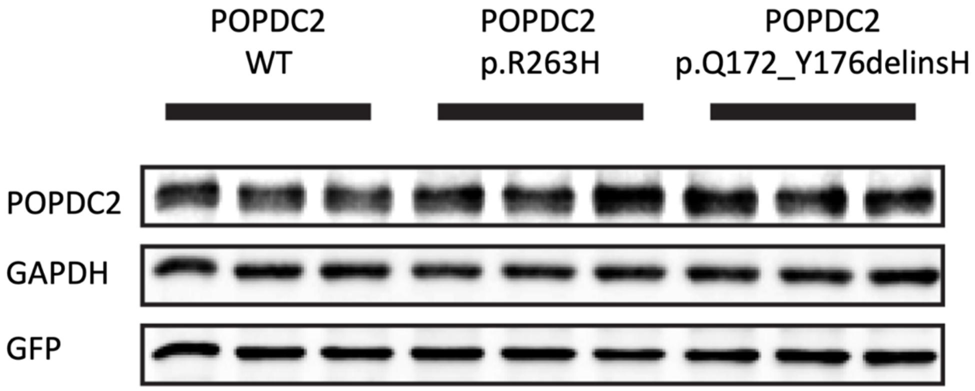

**Supplemental Figure 5.** Western blot analysis of POPDC1 and POPDC2 in protein lysates from muscle biopsies obtained in patient 1 (PT, Subject II-3 from Family A) and in three control subjects (CTRL). Actinin (ACTININ) levels were used as loading control.

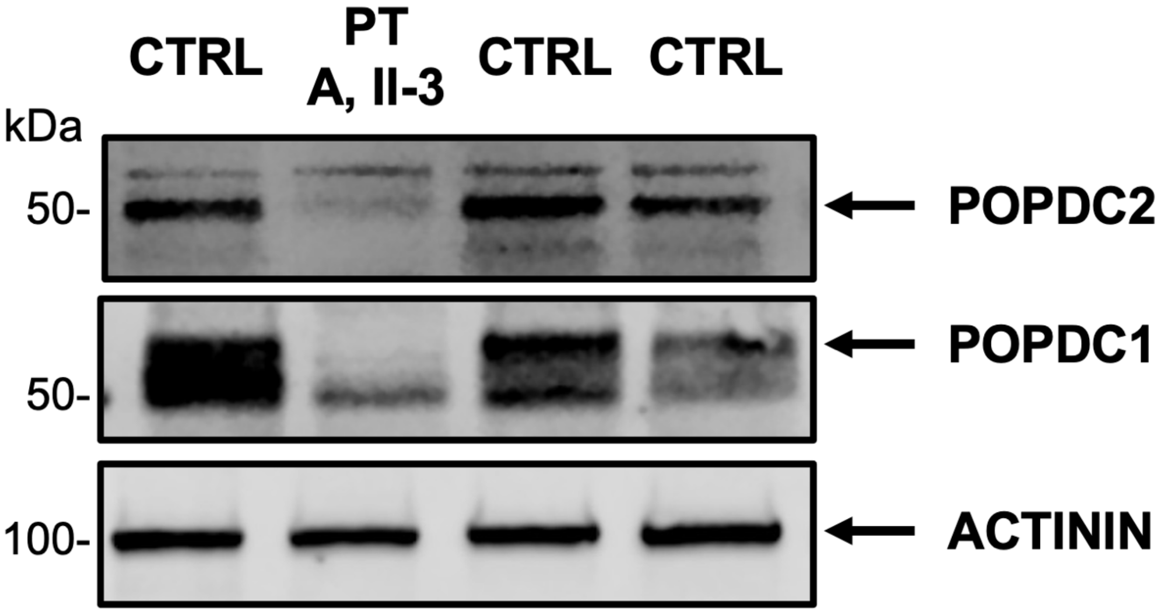

**Supplemental Figure 6.** Expression of Popdc proteins 1–3 in the developing mouse heart. t-SNE maps with a perplexity of 50 were generated for sinus node and atrioventricular node/His single cell RNA-sequencing data obtained from a recently published study.[13] (**A**) Popdc1 (POPDC1), (**B**) Popdc2 (POPDC2) and (**C**) Popdc3 (POPDC3) gene expression intensities were plotted on the t-SNE maps to identify their expression profiles across the present tissue clusters. **Abbreviations:** AV, atrioventricular; CMs, cardiomyocytes; SAN, sinoatrial node; T-SNE, t -distributed stochastic neighbour embedding.

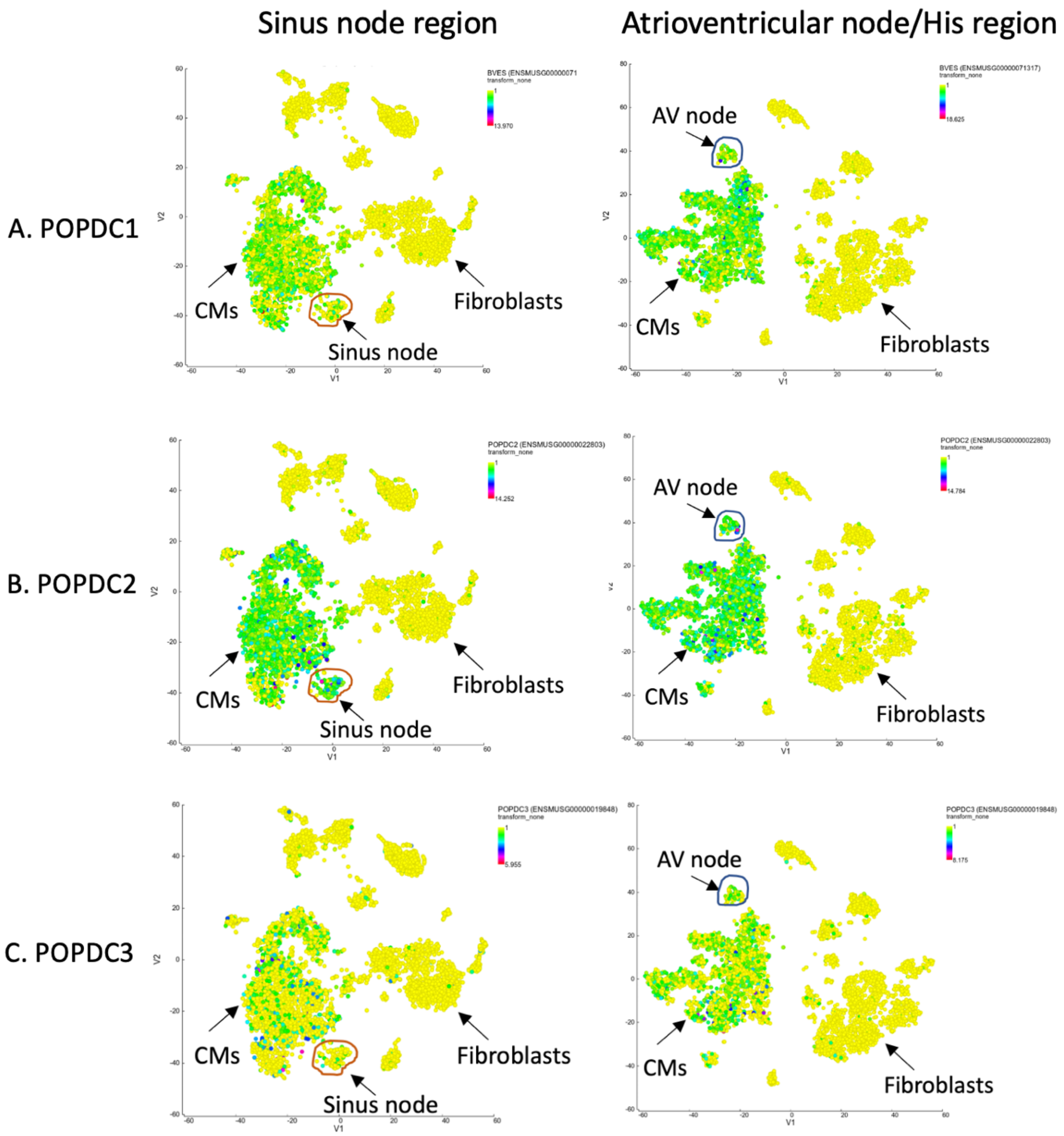
